# Investigating mobility-based fast food outlet visits as indicators of dietary intake and diet-related disease

**DOI:** 10.1101/2021.10.28.21265634

**Authors:** Abigail L. Horn, Brooke M. Bell, Bernardo Garcia Bulle Bueno, Mohsen Bahrami, Burcin Bozkaya, Yan Cui, John P. Wilson, Alex Pentland, Esteban Moro, Kayla de la Haye

## Abstract

**Importance:** Excessive consumption of fast food (FF) is associated with chronic disease. Population-level research on FF outlet visits is now possible with mobility data, however its usefulness as an indicator of FF intake and diet-related disease must be established.

**Objective:** Investigate whether FF outlet visits from mobility data are indicators of self-reported FF intake, obesity, and diabetes, and compared with self-reported intake, equivalent or better indicators of obesity and diabetes.

**Design, Setting, and Participants:** A secondary analysis of data from a representative sample of 8,036 adult residents of Los Angeles County (LAC) from the 2011 Los Angeles County Health Survey (LACHS), and mobility data representing all geolocations between October 2016 - March 2017 of 243,644 anonymous and opted-in smartphone users in LAC.

**Main Outcomes and Measures:** Main outcomes were self-reported FF intake frequency (never, infrequent, moderate, frequent), obesity, and diabetes from LACHS. FF outlet visits were computed as the temporal frequency of FF visits (FF visits/time) and the ratio of visits to FF over all food outlets (FF visits/food), summarized over smartphone users in a neighborhood, scaled from 0-10, and linked to LACHS respondents by census tract.

**Results:** The analytic sample included 5,447 LACHS respondents and 234,995 smartphone users with 14,498,850 visits to food outlets. FF outlet visits were significantly associated with self-reported FF intake (reference: never) for both FF visits/time (infrequent: odds ratio [OR], 1.13; 95% CI, 1.06-1.20; frequent: OR, 1.35; 95% CI, 1.28-1.42) and FF visits/food (infrequent: OR, 1.12; 95% CI, 1.06-1.17; frequent: OR, 1.28; 95% CI, 1.22-1.33). FF outlet visits were significantly associated with obesity (FF visits/time: adjusted OR [AOR], 1.16; 95% CI, 1.12-1.21; FF visits/food: AOR, 1.13; 95% CI, 1.10-1.17) and diabetes (FF visits/time: AOR, 1.15; 95% CI, 1.09-1.21; FF visits/food: AOR, 1.11; 95% CI, 1.07-1.16), adjusted for sociodemographic factors. Models of the association between FF outlet visits and obesity or diabetes had better fits than between self-reported FF intake and obesity or diabetes.

**Conclusions and relevance:** This study illustrates that population-scale mobility data provide useful, passively-collected indicators of FF intake and diet-related disease within large, diverse urban populations that may be better than self-report intake.

**Key Points:** *Question:* Do visits to fast food outlets observed in mobility data provide meaningful measures of fast food intake, and when compared with self-reported intake, equivalent or better indicators of diet-related disease?

*Findings:* In this cross-sectional Los Angeles County study from a survey of 8,036 adults and mobility data from 243,644 smartphone users with 14.5 million food outlet visits, neighborhood-level features representing visits to fast food outlets were significantly associated with self-reported fast food intake, significantly associated with obesity and diabetes, and were a better predictor of these diseases than self-reported fast food intake.

*Meaning:* Measures of food behaviors observed in population-scale mobility data can provide meaningful indicators of food intake and diet-related diseases, and could complement existing dietary surveillance methods.

## Introduction

Food environments, where people acquire and consume food, impact diet and related diseases (i.e., nutritional health)^1^. To date, research has focused on predefined local and *static* food environments, largely of the home neighborhood^2,3^. Their features (e.g., the availability of fast food outlets) can predict nutritional health^1^ although findings are mixed^4–6^. A growing proportion of food acquisition occurs miles from our homes^7^, therefore the limited focus on static food environments may be one cause of these mixed results.

A major gap in the literature is evidence of the *dynamic* food environments people are exposed to in their daily routines (i.e., their “activity space”^8^), the food outlets they visit, and how these mobile food environments impact dietary intake and health. With the availability of big data on human mobility (i.e., geolocations captured by people’s smartphones), population-level research on the food outlets that people have access to and visit given their daily movements is now possible. Some studies (often *n*<100) have begun to use GPS tracking technologies to continuously observe how people navigate their environment to acquire food over relatively brief time periods (i.e., 1 week)^9,10^. However, to our knowledge, *large-scale* mobility data has not been used to study food environments and their connection with nutritional health.

This study undertakes a critical first step in this line of research: investigating whether visits to food outlets observed in population-level mobility data provide meaningful indicators of dietary intake and diet-related disease. We focus these analyses on *fast food* (FF) outlets specifically because FF intake is linked to disease risk^11^, makes up 16% of Americans’ caloric intake^7^, and because FF outlets are well-distributed across food environments.

We utilize a large mobility data set from Los Angeles County (LAC), U.S.A., to generate neighborhood-level measures of visits to FF outlets. The first objective was to determine whether visits to FF outlets from population mobility data are a meaningful indicator of individuals’ self-reported FF intake. The second objective was to determine whether visits to FF outlets (mobility data) are a meaningful predictor of individuals’ obesity and diabetes, and a comparable or better predictor than self-reported FF intake.

## Methods

### Individual Health and Demographic Data Source and Measures

Individual-level measures of FF intake and diet-related disease come from the 2011 Los Angeles County Health Survey (LACHS). LACHS is a population-based dual frame (landline and cellular) telephone survey conducted by Los Angeles County Department of Public Health (LACDPH). It collects data from representative samples of adults and children living within LAC, on topics such as health conditions and behaviors, sociodemographics, and home residence. Our study uses data from the Adult Survey module, which includes 8,036 randomly selected LAC residents who are 18 years and over. Detailed study protocols are available from LACHS^12^.

For this study, we excluded participants who: were missing residential census tract information, lived in a rural census tract^13^, or had missing data for all outcome variables. The final analytic sample was 5,447 participants residing in 1,941 census tracts.

All variables analyzed in this study were self-reported, and some were recoded from the original measures (eMethods the **Supplement**) for ease of interpretability. **FF intake frequency** was coded as a four-category variable: never, infrequent (< once per month), moderate (≥ once per month to < once per week), and frequent (≥ once per week). **Obesity** (having a Body Mass Index, BMI ≥30) and **diabetes** (having a diagnosis) were coded as binary variables (yes/no). **Sociodemographic factors** included age group, gender, race/ethnicity, educational level, and household income level. Respondents’ census tract of residence was derived from their home address.

### Geolocation (Mobility) Data Source and Measures

Geolocation (i.e., mobility) data were collected by Cuebiq^14^, a location-based services company that maintains anonymized geospatial datasets on human mobility by aggregating data across smartphone applications from mobile phone devices. The dataset consists of anonymized records of GPS locations from individual adult (≥18) smartphone users who have opted in to provide access to their GPS location data anonymously through a General Data Protection Regulation and California Consumer Privacy Act compliant framework. Users across all major smartphone device operating systems (e.g., iOS, Android, Windows) are represented. The dataset includes 243,644 users with estimated residential census tracts (explained below) in LAC between October 2016 - March 2017 (6 months), representing 3.1% of the LAC adult population^15^.

The data consist of geolocation “pings” identifying the location of a given smartphone, typically recorded every 5-15 minutes (eFigure 1 in the **Supplement**). Each ping contains the GPS location of the phone (latitude and longitude), timestamp, and anonymous (encrypted and hashed) identifier which is unique for all smartphone users. No other individual information (e.g., demographic characteristics) was available on users. From the trajectories of pings for each user, we used a detection algorithm^16^ to filter out transient locations and extract meaningful “stays” (or stops) at particular locations of at least 5 minutes duration. We excluded users if they had fewer than two stays at any location over the 6 months, resulting in an analytic sample of 234,995 users with over 63 million observed stays (eTable 1 in the **Supplement**).

Visits to food and FF outlets were identified by linking geolocated stays with a points of interest (POI) database obtained from the public Foursquare API^17^ in 2017, which provides the names and geolocations of 239,509 POI in LAC. Food outlets were defined as any location where food might be sold (including restaurants, food retailers, and other locations) and FF outlets were defined as limited-service restaurants serving menus of predominantly ultra-processed and/or low-nutrient, energy dense foods (e.g., McDonald’s, Taco Bell, Pizza Hut). We identified food and FF outlets using a combination of Foursquare’s existing categorization taxonomy, and a bottom-up search of known chain FF outlet names validated in previous research (eMethods and eTables 2-3 in the **Supplement**)^18,19^. After recoding, there were 53,588 food outlets and 4,151 FF outlets. A total of 14,498,850 visits to food outlets were detected across the analytic sample.

We estimated the home residential census tract for each user as the tract in which the majority of their activity between 10pm-6am occurred. Using the preprocessed mobility data, FF outlet visits were defined first at the level of an individual user, and then aggregated and averaged across users living within 247 LAC neighborhoods^20^ to represent the “typical” FF visit behavior of residents in that spatial area. The neighborhood level was the smallest administrative area we could demonstrate that our mobility user sample achieved broad geographic representation of the underlying population (eMethods and eFigures 4-6 in the **Supplement**).

The first FF outlet visit variable, the **temporal frequency of FF outlet visits (FF visits/time)**, was defined as the percentage of observed periods (out of three possible daily periods: before 11am, 11am-4pm, after 4pm) in which a user visits at least one FF outlet, out of the total number of observed periods for that user. The second variable, **relative frequency of FF to all food outlet visits (FF visits/food)**, was defined as the percentage of the total number of visits to FF outlets for a user out of the total number of visits to any food outlet for that user. We also defined a covariate representing **average mobility behavior**, as the average number of trips, measured as “stays” (defined above) per user per day (**trips/day**). These three variables were averaged over all users with an estimated home residence within a neighborhood, rescaled from 0-10 to enable comparison of effect sizes in regression analyses, and linked as contextual variables to individual respondents from the LACHS survey based on their home census tract of residence. A geographic visualization of the unscaled mobility variables is provided in the **Figure**.

We used sample post-stratification techniques, comparison with census data, and sensitivity tests on the mobility data to establish (i) its measurement accuracy, (ii) the validity of approaches taken to attribute stays to POI, and (iii) its population representativeness at the neighborhood level; see eMethods, eTables 4-5, and eFigures 3-6 in the **Supplement**.

All study protocols were approved by the Institutional Review Boards (IRBs) of the LACDPH, University of Southern California, and Massachusetts Institute of Technology. Where applicable, this study followed the Strengthening the Reporting of Observational Studies in Epidemiology (STROBE) reporting guidelines.

### Statistical Analysis

Logistic regression models at the LACHS respondent-level with linked mobility variables were generated to test the study objectives. For the first objective, unadjusted (univariable) and adjusted (multivariable) multinomial logistic regression models were used to estimate the odds ratios (ORs) for the association between the two FF outlet visit variables (IV), and self-reported FF intake frequency as the dependent variable (DV). The multivariable models adjusted for sociodemographic factors. *P-*values for the statistical significance of differences between adjusted and unadjusted ORs were tested using a χ^2^ test.

For the second objective, multivariable logistic regression models tested whether (i) FF visits/time (IV), (ii) FF visits/food (IV), and (iii) self-reported FF intake (IV) were associated with obesity (DV), and in separate models, with diabetes (DV). All models adjusted for sociodemographic factors. ORs for the categorical variable (FF intake frequency) and continuous variables (FF visits/time and FF visits/food) cannot be directly compared. Therefore, model fits were compared on the basis of their Akaike weights, which are transformations from raw Akaike information criterion (AIC) values to facilitate interpretation of AIC model comparisons^21^. A model’s Akaike weight is interpreted as its probability of being the best out of a set of candidate models.

The LACHS and mobility datasets are from different years because FF intake was not assessed by the LACHS survey after 2011, while geolocation-based mobility data was not available before 2016. We conducted sensitivity analyses to examine whether regression model results were impacted by this time gap. Sensitivity analyses were also conducted to examine whether results were impacted by controlling for users’ general mobility behavior (Trips/day). Mobility data were analyzed in Python. LACHS data and statistical analyses were conducted using R software, version 3.6.3. A 2-sided *P* < .05 was considered statistically significant. Additional data and analysis details are available in eMethods in the **Supplement**.

## Results

### Descriptive Results

When comparing the full (n=8,036) and analytic (n=5,447) samples of LACHS respondents, we found small (1-3%) but statistically significant differences in age group, gender, race/ethnicity, household income level, and self-reported FF intake frequency (eTable 6 in the **Supplement**). The analytic sample had representation across all age groups, race/ethnicity groups, and education levels; and had higher proportions of female vs. male (**Table 1**). Of the analytic sample, 17.3% reported never eating FF, 19.1% reported infrequent intake, 26.9% reported moderate intake, and 36.7% reported frequent intake; 24.8% had obesity; and 11.1% had diabetes.

**Table 1.**
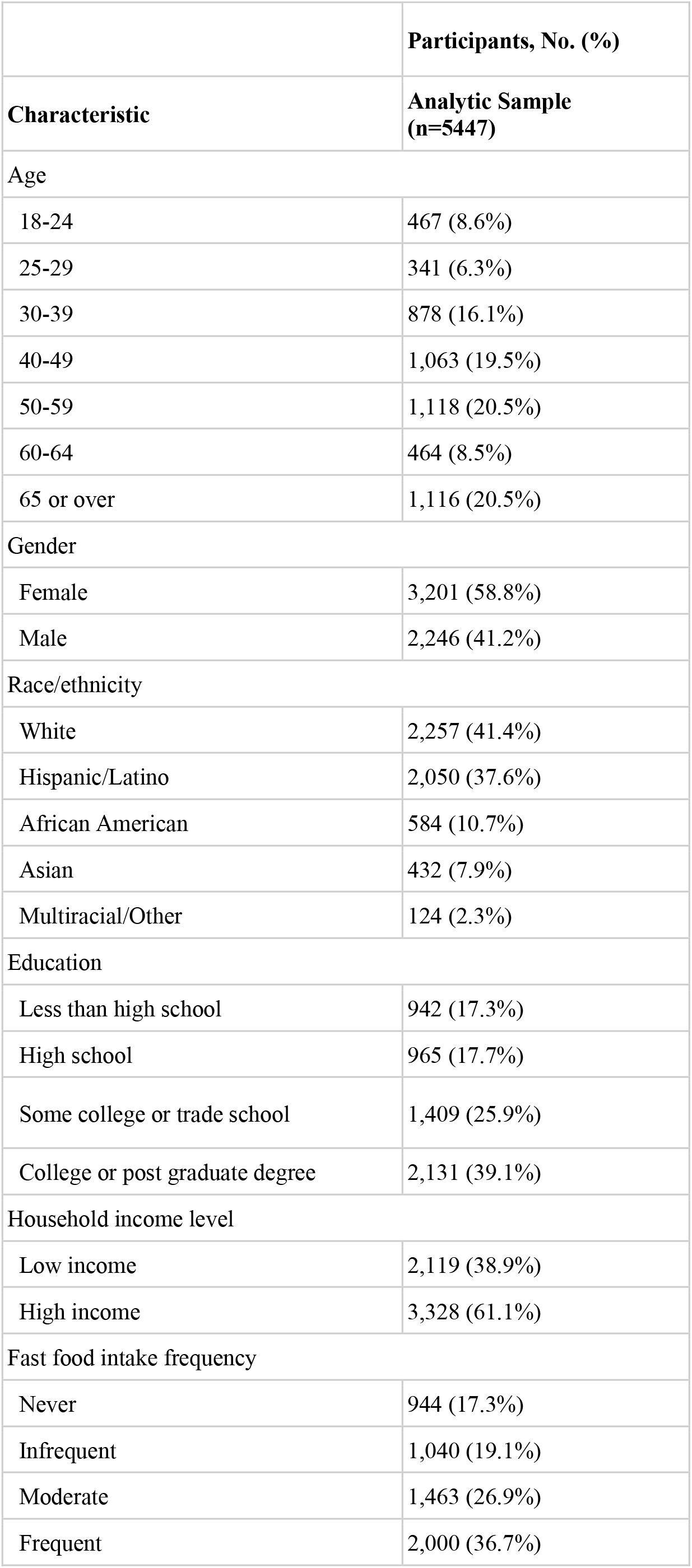
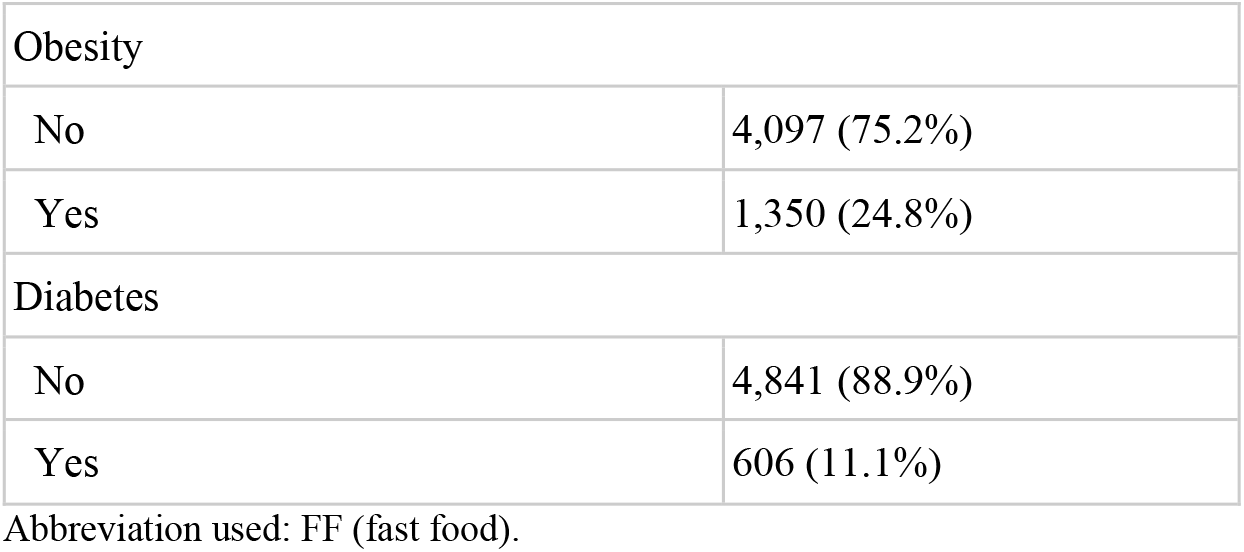
Demographic, Diet, and Diet-related Disease Characteristics For Analytic Sample of LACHS Participants.

Across the mobility variables linked to LACHS respondents, the median percentage of observed daily periods in which users visited FF outlets (FF visits/time unscaled) was 4.3% (range, 1.0-13.0%); the median percentage of visits to all food outlets that were FF (FF visits/food unscaled) was 10.6% (range, 2.8-22.4%); and the median number of trips per day (trips/day unscaled) was 4.0 (range, 2.2-8.0) (see eFigure 7 in the **Supplement** for histograms of these distributions).

### Association Between Visits to FF Outlets and FF Intake Frequency

In the unadjusted models, higher frequencies of FF outlet visits were significantly associated with higher levels of FF intake for both variables (**Table 2**). For a 10% increase in FF visits/time (i.e., a 1-unit increase in the scaled variable), relative to no FF intake, the odds of high FF intake increased by 35% (OR, 1.35; 95% CI, 1.28-1.42), of moderate FF intake increased by 26% (OR, 1.26; 95% CI, 1.19-1.33), and of low FF intake increased by 13% (OR, 1.13; 95% CI, 1.06-1.2). For a 10% increase in the frequency of FF visits/food, relative to no FF intake, the odds of high FF intake increased by 28% (OR, 1.28; 95% CI, 1.22-1.33), of moderate FF intake increased by 22% (OR, 1.22; 95% CI, 1.16-1.27), and of infrequent FF intake increased by 12% (OR, 1.12; 95% CI, 1.06-1.17).

**Table 2.** Odds Ratios for Unadjusted and Adjusted Multinomial Logistic Regression Analyses of the Association Between Visits to Fast Food Outlets Observed in Mobility Data and Self-Reported Fast Food Intake^a^.

|  | Unadjusted analysis |  |  | Adjusted analysis<sup>c</sup> |  |  |
| --- | --- | --- | --- | --- | --- | --- |
| Model<sup>b</sup> | FF intake frequency, OR (95% CI) |  |  | FF intake frequency, OR (95% CI) |  |  |
|  | Infrequent | Moderate | Frequent | Infrequent | Moderate | Frequent |
| FF visits/time | 1.13 (1.06-1.20) | 1.26 (1.19-1.33) | 1.35 (1.28-1.42) | 1.12 (1.05-1.20) | 1.22 (1.15-1.30) | 1.30 (1.23-1.38) |
| FF visits/food | 1.12 (1.06-1.17) | 1.22 (1.16-1.27) | 1.28 (1.22-1.33) | 1.11 (1.06-1.17) | 1.19 (1.14-1.24) | 1.25 (1.19-1.31) |

Abbreviations: OR, odds ratio; CI, confidence interval; FF, fast food.
<sup>a</sup> Multinomial logistic regression models for fast food intake frequency across four frequency categories. Reference group: no fast food intake. $P < .001$ for each estimated odds ratio.
<sup>b</sup> Each model estimated fast food intake frequency using the fast food visit variable listed in this column as the primary independent variable.
<sup>c</sup> Adjusted for age group, gender, race/ethnicity, educational level, and household income level.
<sup>d</sup> $P = .01$ .

In the adjusted models, all three measures of FF outlet visits remained strong and significant predictors of FF intake (**Table 2**), and the estimates were essentially unchanged; the CIs of the adjusted vs. unadjusted ORs are overlapping and there were no significant differences between *P* values (eTable 7 in the **Supplement**).

### Association Between Visits to FF Outlets and Diet-Related Disease

A 10% increase in FF visits/time was significantly associated with a 16% greater odds of obesity (AOR, 1.16; 95% CI, 1.12-1.21) and a 15% greater odds of diabetes (AOR, 1.15; 95% CI, 1.09-1.21) (**Table 3**). A 10% increase in FF visits/food was significantly associated with a 13% greater odds of obesity (AOR, 1.13; 95% CI, 1.10-1.17) and an 11% greater odds of diabetes (AOR, 1.11; 95% CI, 1.07-1.16). Self-reported infrequent FF intake frequencies were not significantly associated with obesity or diabetes, while moderate FF intake was significantly associated with obesity (AOR, 1.26; 95% CI, 1.03-1.55), and frequent FF intake was significantly associated with obesity (AOR, 1.63; 95% CI, 1.34-1.99) and diabetes (AOR, 1.39; 95% CI, 1.08-1.81).

**Table 3.**
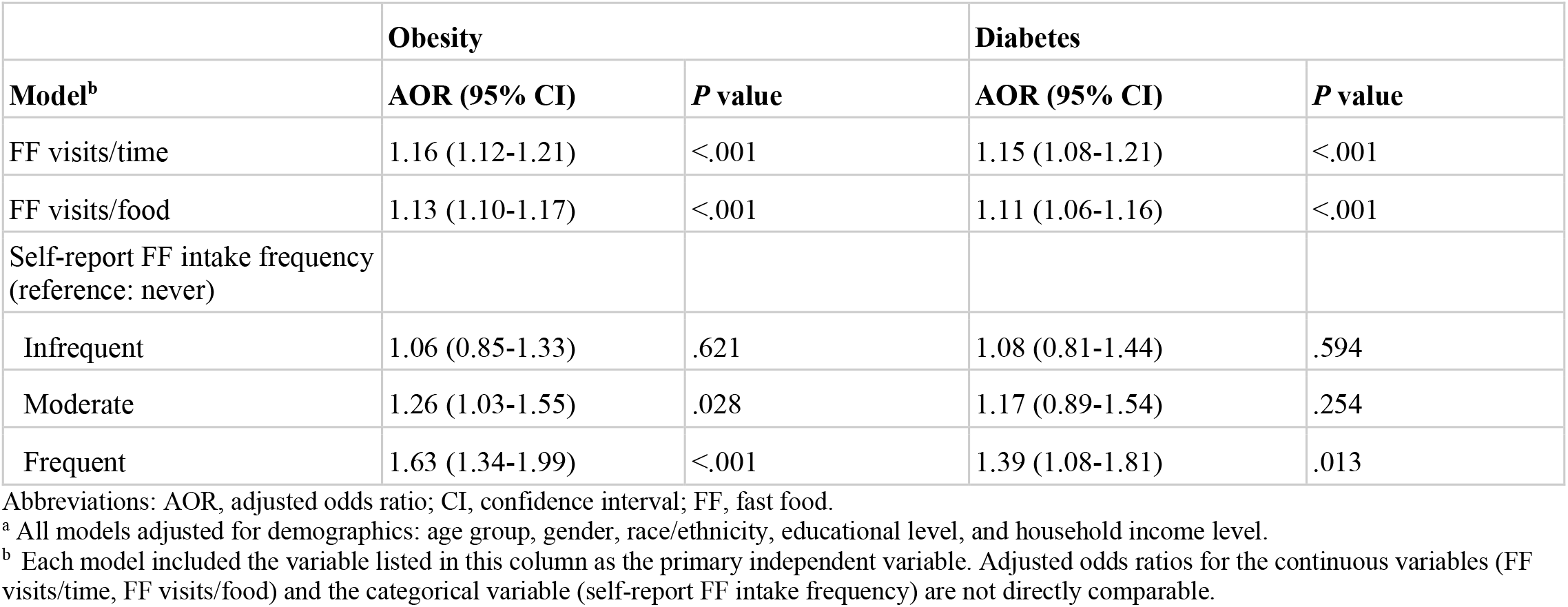
Odds Ratios for Multivariable Binary Logistic Regression Analyses of the Association Between Visits to Fast Food Outlets and Diet-Related Disease^a^.

|  | **Obesity** |  | **Diabetes** |  |
| --- | --- | --- | --- | --- |
| **Model<sup>b</sup>** | **AOR (95% CI)** | **P value** | **AOR (95% CI)** | **P value** |
| FF visits/time | 1.16 (1.12-1.21) | <.001 | 1.15 (1.08-1.21) | <.001 |
| FF visits/food | 1.13 (1.10-1.17) | <.001 | 1.11 (1.06-1.16) | <.001 |
| Self-report FF intake frequency (reference: never) |  |  |  |  |
| Infrequent | 1.06 (0.85-1.33) | .621 | 1.08 (0.81-1.44) | .594 |
| Moderate | 1.26 (1.03-1.55) | .028 | 1.17 (0.89-1.54) | .254 |
| Frequent | 1.63 (1.34-1.99) | <.001 | 1.39 (1.08-1.81) | .013 |

Abbreviations: AOR, adjusted odds ratio; CI, confidence interval; FF, fast food.
<sup>a</sup> All models adjusted for demographics: age group, gender, race/ethnicity, educational level, and household income level.
<sup>b</sup> Each model included the variable listed in this column as the primary independent variable. Adjusted odds ratios for the continuous variables (FF visits/time, FF visits/food) and the categorical variable (self-report FF intake frequency) are not directly comparable.

Comparing Akaike weights across the models of obesity, the probability that the model including FF visits/time was the best-fitting model was 0.10, including FF visits/food was 0.90, and including FF intake frequency was 3.6e-7 (**Table 4**). Comparing Akaike weights across the three models of diabetes, the probability that the model including FF visits/time was the best-fitting model was 0.69, including FF visits/food was 0.31, and including self-reported FF intake frequency was 2.3e-5.

**Table 4.** Values of the Akaike Information Criterion (AIC) and Akaike Weights Calculated From Models of the Association Between Visits to Fast Food Outlets, Self-Report Fast Food Intake Frequency, and Diet-Related Disease^a^.

|  | **Obesity** |  | **Diabetes** |  |
| --- | --- | --- | --- | --- |
| **Model** | **AIC** | **Akaike weight<sup>b</sup>** | **AIC** | **Akaike weight** |
| FF visits/time | 5754.7 | .10 | 3353.8 | .69 |
| FF visits/food | 5750.2 | .90 | 3355.4 | .31 |
| Self-report FF intake frequency | 5777.9 | 3.6e-7 | 3374.4 | 2.3e-5 |

Abbreviations: AIC, Akaike information criterion; FF, fast food.
<sup>a</sup> All models adjusted for demographics: age group, gender, race/ethnicity, educational level, and household income level. Each model included the variable listed in this column as the primary independent variable alongside demographic covariates.
<sup>b</sup> Each model's Akaike weight can be interpreted as the probability that it is the best model out of the set of three candidate models.

Regression results were not significantly impacted by (i) controlling for general mobility behavior (Trips/day) (eTable 8 in the **Supplement**), and (ii) the time gap between data collection for the LACHS (2011) and mobility data (2016-17) (eMethods, eTables 9-14, and eFigure 8 in the **Supplement**).

## Discussion

Using large-scale mobility data from LAC, this study finds strong and consistent evidence that visits to FF outlets, aggregated at the neighborhood level, strongly and significantly correspond to individuals’ consumption of FF. Thus, passively observed visits to FF outlets appear to be a good indicator of FF intake in a diverse, urban population.

FF behaviors observed in the mobility data also predicted diet-related disease. Moreover, models of the association between FF outlet visits and obesity or diabetes had substantially *better* fits than between self-reported FF intake, the standard measure in population nutrition research, and obesity or diabetes. Findings held after controlling for individual sociodemographic factors and general mobility behavior, suggesting these indicators are uniquely representing visits to FF outlets rather than mobility alone. Several factors may explain the strength of this result, despite aggregation at the neighborhood level. First, measures of food behaviors observed directly from smartphone-captured mobility data may be less prone to measurement error compared with self-reported food intake. This may be related to mobility data capturing behavior continuously, over months (or potentially longer), recording more detail of behavioral patterns than can be reliably assessed by self-reported recall^22,23^. Second, the neighborhood-aggregated measures of visits to FF outlets may be capturing an indicator of social behaviors of a larger group. Because eating is strongly influenced by social and cultural factors^24^, the ‘social signal’^25^ captured by the aggregated measure may be additionally predictive of disease risk. Third, the indicators may be picking up other neighborhood-level risk factors for these diseases.

This study advances research methods that are used to understand how food environments are related to health. It establishes that features extracted from large-scale mobility data provide a strong signal of FF consumption, suggesting that this data source may represent a valid population surveillance tool for this and possibly other eating and health behaviors. Mobility data is objective, and captured passively and continuously over long periods of time, making it a convenient and information-rich means of gathering population-level food behaviors that are notoriously hard to measure.

More broadly, this study introduces human mobility data as an untapped resource for future investigations into links between food environment use and nutritional health across large, diverse populations. Studies involving mobility data might include: re-defining notions of “food deserts” and “food swamps” to account for lived environments beyond the home neighborhood; investigating routine behaviors that determine spatio-temporal accessibility to different types of food environments; using “natural experiments” (e.g., users who change home or food environments) to identify causal mechanisms linking features of food environments and eating behaviors; and developing more effective policies and interventions on food environments that take routine behaviors beyond the home neighborhood into consideration.

### Limitations

The results of this study are prone to ecological fallacy, since measures of FF outlet visits averaged across the aggregate of individuals within a neighborhood group are assumed to apply to all individuals within that group. Summarizing mobility features at the neighborhood (or other spatial) level will obscure group differences, for example, differential visits to FF outlets based on gender or other demographics^26^. Additionally, the mobility measures represent sample means over a convenience sample of mobility users living in a neighborhood, which may not reflect the true mean within that neighborhood.

Smartphone users, although constituting 83% of the U.S. adult population in 2017^27^, represent a subset of the population that has some uneven representation across socio-demographic groups (e.g., low income^27^, older and non-white^28^), leading to potential biases. Quantifying these biases in mobility data is challenging since demographic information is not available on individual smartphone users, however we use post-stratification sampling on the mobility data to achieve population representativeness at the level of the neighborhoods, which may alleviate some of the potential biases (eMethods in the **Supplement**). Additionally, our previous work on this dataset demonstrated low bias across income classes by imputing this characteristic for each user^29^. Different study designs will be necessary to investigate whether inferences derived from smartphone users are fully generalizable to these populations.

We have linked two different study populations and timeframes, which leads to potential sources of incompatibility. There may have been changes in food environments between 2011 and 2016-17 that could affect population visits to food outlets. However, we found that there has been very little change in population demographics in the LAC study areas between 2011 and 2017, and that regression model results were robust to removing the census tracts with the greatest change (eMethods and eTables 9-14 in the **Supplement**).

Limitations of the mobility data include under-sampled visits to FF outlets due to the gaps in measurement of each smartphone user (e.g., when phones are out of service). We address this by defining percentage-based variables in which observations of FF outlet visits are relativized to other observables (e.g., all food outlet visits), but there may be measurement error that cannot be quantified. Separately, the FF intake measures may be subject to biases common to self-report food frequency measures.

While we have taken several approaches to validate the measurement accuracy of the mobility data and robustness of findings to our methods for attributing geolocations to places, there may be limitations to our ability to detect visits to certain food outlets (eMethods in the **Supplement**).

## Conclusions

Population-scale mobility data provide rich information about population use of fast food (FF) outlets. While there are sources of bias in mobility data, this study demonstrates that it provides (i) useful indicators of FF intake within large and diverse urban populations and (ii) meaningful predictors of diet-related disease (i.e., obesity and diabetes), and may have advantages over existing dietary assessment methods. Mobility data are likely to facilitate future research investigating how people of diverse backgrounds move around to dynamically use food environments, including and beyond the home neighborhood, and the links to their diet and health.

## Supporting information

Supplementary Online Content

## Data Availability

Two datasets were analyzed in this manuscript. Individual-level measures of FF intake and diet-related disease analyzed in this study came from the 2011 Los Angeles County Health Survey (LACHS). This data was shared privately under an IRB-approved Data Use Agreement with the University of Southern California. These data cannot be shared.
Geolocation (i.e., mobility) data were collected by Cuebiq Inc., consisting of anonymized records of GPS locations from individual adult (≥18) smartphone users who have opted in to provide access to their GPS location data anonymously through a General Data Protection Regulation and California Consumer Privacy Act compliant framework, were collected by Cuebiq Inc. and shared with the Massachusetts Institute of Technology through an IRB-approved Data License Agreement via their Data for Good Program. These data cannot be shared.

## Author Contributions

Dr. Horn had full access to all the data in the study and takes responsibility for the integrity of the data and the accuracy of the data analysis.

*Concept and design*: Horn, Bozkaya, Moro, de la Haye.

*Acquisition, analysis, or interpretation of data*: Horn, Bell, Bulle Bueno, Bahrami, Cui, Wilson, Pentland, Moro, de la Haye.

*Drafting of the manuscript:* Horn, Bell, de la Haye.

*Critical revision of the manuscript for important intellectual content*: All authors.

*Statistical analysis*: Horn, Bell, Bulle Bueno.

*Obtained funding*: Horn, Moro, de la Haye.

*Supervision*: Moro, de la Haye.

## Conflict of Interest Disclosures

None reported.

## Funding/Support

This work was supported with funding from the University of Southern California’s Keck School of Medicine Dean’s Pilot Funding Program (PI: de la Haye) and by MIT Connection Science (PIs: Pentland and Moro). Dr. Horn acknowledges support from an NIH Ruth L Kirschstein National 591 Research Service Award (NRSA) Institutional Training Grant (T32 5T32CA009492-35). Dr. Moro acknowledges funding from Agencia Estatal de Investigación (Spain) (PID2019-106811GB-C32/AEI/10.13039/501100011033)

## Role of the Funder/Sponsor

The funder/sponsor had no role in the design and conduct of the study; collection, management, analysis, and interpretation of the data; preparation, review, or approval of the manuscript; and decision to submit the manuscript for publication.

## Additional Contributions

Cuebiq Inc. shared the data through an IRB-approved Data License Agreement via their Data for Good Program and approved the final version of the manuscript. Ashlesha Datar, PhD, shared a list of fast food outlet brands. None were compensated for their work.

---

**Figure 1.**
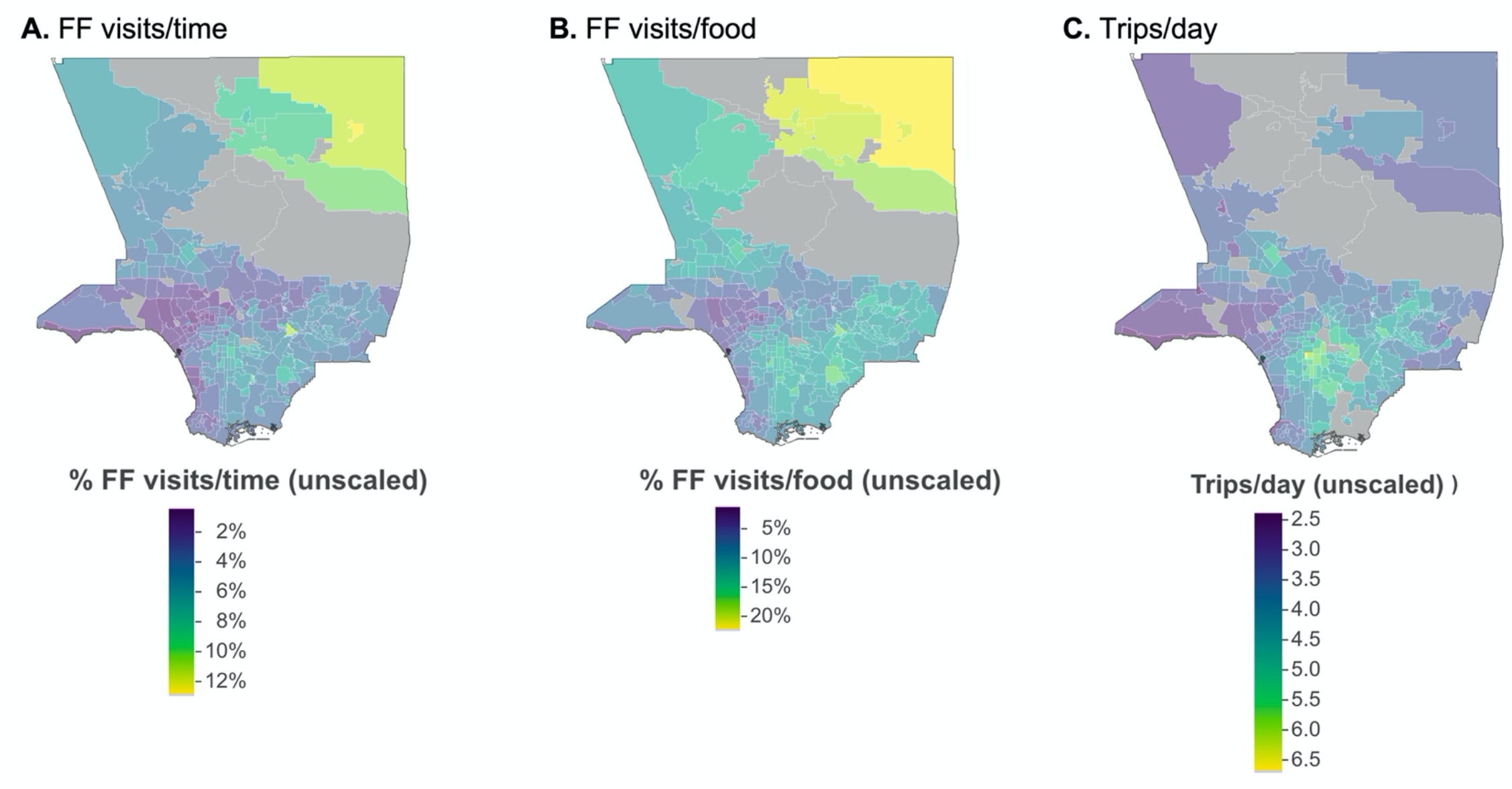
Geographic Distribution of the Unscaled Mobility Variables Across Los Angeles County Neighborhoods. Abbreviations: LACHS, Los Angeles County Health Survey; FF, fast food. Figure represents the geographical distribution of the unscaled mobility variables across the 247 neighborhoods with mobility users having an estimated home address in the neighborhood, before linkage to LACHS respondents. Areas with gray shading represent neighborhoods not included in the study because of either no residing mobility users, or because they contained predominantly rural census tracts^13^. FF visits/time unscaled (A) represents the percentage of observed daily periods in which users visited FF outlets. FF visits/food unscaled (B) represents the percentage of visits to all food outlets that were FF. Trips / day unscaled (C) represents the number of trips per day across all users within a neighborhood. Mobility variables are averaged over all users with an estimated home residence within a neighborhood.

## Notes

### Competing Interest Statement

The authors have declared no competing interest.

### Author Declarations

All study protocols were approved by the Institutional Review Boards (IRBs) of (i) the Los Angeles County Department of Public Health, (ii) the University of Southern California, and (iii) the Massachusetts Institute of Technology.

## References

1. Story M, Kaphingst KM, Robinson-O’Brien R, Glanz K. Creating Healthy Food and Eating Environments: Policy and Environmental Approaches. Annu Rev Public Health. 2008;29(1):253–272. doi:10.1146/annurev.publhealth.29.020907.090926

2. Feng J, Glass T, Curriero F, Stewart W, Schwartz B. The built environment and obesity: a systematic review of the epidemiologic evidence. Health Place. 2010;16(2):175–190. doi:10.1016/J.HEALTHPLACE.2009.09.008

3. Leal C, Chaix B. The influence of geographic life environments on cardiometabolic risk factors: a systematic review, a methodological assessment and a research agenda. Obes Rev. 2011;12(3):217–230. doi:10.1111/J.1467-789X.2010.00726.X

4. Dubowitz T, Zenk SN, Ghosh-Dastidar B, et al. Healthy food access for urban food desert residents: examination of the food environment, food purchasing practices, diet and BMI. Published online 2014. doi:10.1017/S1368980014002742

5. Cobb L, Appel L, Franco M, Jones-Smith J, Nur A, Anderson C. The relationship of the local food environment with obesity: A systematic review of methods, study quality, and results. Obesity (Silver Spring). 2015;23(7):1331–1344. doi:10.1002/OBY.21118

6. Chen X, Kwan MP. Contextual uncertainties, human mobility, and perceived food environment: The uncertain geographic context problem in food access research. Am J Public Health. 2015;105(9). doi:10.2105/AJPH.2015.302792

7. Saksena MJ, Okrent AM, Anekwe TD, et al. America’s Eating Habits: Food Away From Home. Published online 2018:EIB-196.

8. James P, Jankowska M, Marx C, et al. Spatial Energetics: Integrating Data From GPS, Accelerometry, and GIS to Address Obesity and Inactivity. Am J Prev Med. 2016;51(5):792–800. doi:10.1016/j.amepre.2016.06.006

9. Chaix B. Mobile Sensing in Environmental Health and Neighborhood Research. Annu Rev Public Heal. 2018;39:367–384. doi:10.1146/annurev-publhealth

10. Scully JY, Moudon AV, Hurvitz PM, Aggarwal A, Drewnowski A. A Time-Based Objective Measure of Exposure to the Food Environment. Int J Environ Res Public Heal 2019, Vol 16, Page 1180. 2019;16(7):1180. doi:10.3390/IJERPH16071180

11. Bahadoran Z, Mirmiran P, Azizi F. Fast Food Pattern and Cardiometabolic Disorders: A Review of Current Studies. Heal Promot Perspect. 2015;5(4):231. doi:10.15171/HPP.2015.028

12. Los Angeles County Department of Public Health. Los Angeles County Health Survey. Accessed October 20, 2021. http://www.publichealth.lacounty.gov/ha/hasurveyintro.htm

13. United States Department of Agriculture Economic Research Service. Rural-Urban Commuting Area Codes (Updated 7/3/2019). Published 2014. Accessed October 20, 2021. https://www.ers.usda.gov/data-products/rural-urban-commuting-area-codes.aspx

14. Cuebiq Data for Good Program. Accessed October 20, 2021. https://www.cuebiq.com/about/data-for-good/

15. United States Census Bureau. 2017 American Community Survey 5-Year Data. Accessed October 20, 2021. https://www.census.gov/programs-surveys/acs

16. Hariharan R, Toyama K. Project Lachesis: Parsing and Modeling Location Histories. In: Springer, Berlin, Heidelberg; 2004:106–124. doi:10.1007/978-3-540-30231-5_8

17. Foursquare API. Accessed October 20, 2021. https://developer.foursquare.com/

18. Datar A, Nicosia N. Assessing social contagion in body mass index, overweight, and obesity using a natural experiment. JAMA Pediatr. 2018;172(3). doi:10.1001/jamapediatrics.2017.4882

19. Datar A, Mahler A, Nicosia N. Association of Exposure to Communities With High Obesity With Body Type Norms and Obesity Risk Among Teenagers. JAMA Netw open. 2020;3(3). doi:10.1001/jamanetworkopen.2020.0846

20. The Los Angeles Times Datadesk. Mapping L.A. Neighborhoods. Accessed October 20, 2021. http://maps.latimes.com/neighborhoods/

21. Wagenmakers EJ, Farrell S. AIC model selection using Akaike weights. Psychon Bull Rev. 2004;11(1). doi:10.3758/BF03206482

22. Riley WT, Rivera DE, Atienza AA, Nilsen W, Allison SM, Mermelstein R. Health behavior models in the age of mobile interventions: Are our theories up to the task? Transl Behav Med. 2011;1(1). doi:10.1007/s13142-011-0021-7

23. Spruijt-Metz D, Hekler E, Saranummi N, et al. Building new computational models to support health behavior change and maintenance: new opportunities in behavioral research. Transl Behav Med. 2015;5(3). doi:10.1007/s13142-015-0324-1

24. Herman CP, Polivy J, Pliner P, Vartanian LR. Social Influences on Eating.; 2019. doi:10.1007/978-3-030-28817-4

25. Galesic M, Bruine de Bruin W, Dalege J, et al. Human social sensing is an untapped resource for computational social science. Nature. 2021;595(7866). doi:10.1038/s41586-021-03649-2

26. Smith LG, Widener MJ, Liu B, et al. Comparing Household and Individual Measures of Access through a Food Environment Lens: What Household Food Opportunities Are Missed When Measuring Access to Food Retail at the Individual Level? Ann Am Assoc Geogr. Published online 2021. doi:10.1080/24694452.2021.1930513

27. Pew Research Center. Share of adults in the United States who owned a smartphone from 2011 to 2017, by location. In Statista - The Statistics Portal. Accessed June 22, 2018. https://www.statista.com/statistics/195003/percentage-of-us-smartphone-owners-by-geographic-location

28. Coston A, Guha N, Ouyang D, Lu L, Chouldechova A, Ho DE. Leveraging administrative data for bias audits: Assessing disparate coverage with mobility data for COVID-19 Policy. In: FAccT 2021 - Proceedings of the 2021 ACM Conference on Fairness, Accountability, and Transparency. ; 2021. doi:10.1145/3442188.3445881

29. Moro E, Calacci D, Dong X, Pentland A. Mobility patterns are associated with experienced income segregation in large US cities. Nat Commun. 2021;12(1). doi:10.1038/s41467-021-24899-8

