## Supplementary Online Content for "Investigating mobility-based fast food outlet visits as indicators of dietary intake and diet-related disease"

**eMethods.** Data and Analysis Methods
**eTable 1.** Statistics of the Distribution of Individual Stays in the Mobility Data Across the Analytic Sample of Smartphone Users
**eTable 2.** Non-Food Primary Category and Subcategory Combinations Coded as *food outlets*
**eTable 3.** List of Fast Food Outlet Names
**eTable 4.** Statistics of Stays at Each Value of Thresholding Maximum Distance
**eTable 5.** Unadjusted Odds Ratios of Self-Reported Fast Food Intake in Robustness Analyses of the Attribution of Stays Detected in Mobility Data to Points of Interest
**eTable 6.** Demographic, Diet, and Diet-related Disease Characteristics in the Full and Analytic Samples of LACHS Participants
**eTable 7.** *P* Value for the Difference Between Unadjusted and Adjusted Odds Ratios From Regression Analyses of Self-Reported Fast Food Intake
**eTable 8.** Logistic Regression Analyses of the Association Between Visits to Fast Food Outlets With Diet-Related Disease Adjusted for General Mobility Behavior
**eTable 9.** Number of Census Tracts and LACHS Respondents Removed After Identifying Outliers in Census Tract Variables Between 2011 and 2017
**eTable 10.** Unadjusted Odds Ratios From Regression Models of Self-Reported Fast Food Intake From Sensitivity Analyses to Time Change Between 2011-2017
**eTable 11.** *P* Values for Differences Between Odds Ratios From Regression Models of Fast Food Intake From Sensitivity Analyses to Time Change Between 2011-2017
**eTable 12.** Adjusted Odds Ratios From Regression Models of Obesity From Sensitivity Analyses to Time Change Between 2011-2017
**eTable 13.** Adjusted Odds Ratios From Regression Models of Diabetes From Sensitivity Analyses to Time Change Between 2011-2017
**eTable 14.** *P* Values for Differences Between Odds Ratios From Regression Models of Obesity and Diabetes From Sensitivity Analyses to Time Change Between 2011-2017
**eFigure 1.** Frequency Distribution of the Accuracy of all Pings Generated By a Random Sample of 2,000 (0.8%) of Smartphone Users in the Mobility Dataset
**eFigure 2.** Map of the Spatial Boundaries of the 272 Los Angeles County Neighborhoods
**eFigure 3.** Pearson Correlation Between the Values of Each FF Outlet Visit Variable at Each Value of the Thresholding Distance Between a Stay and POI
**eFigure 4.** Distribution of the Number of Smartphone Users in Analytic Sample Within Each Area for the Census Tract and Neighborhood Spatial Levels
**eFigure 5:** Correlation Between the Smartphone Population Detected in Mobility Data and Census Population at the Census Tract vs. Neighborhood Levels
**eFigure 6.** Correlation Between Post-Stratified (Weighted) and Unweighted Fast Food Outlet Visit Variables
**eFigure 7.** Histograms of the Distribution of the Mobility Variables Linked to LACHS Respondents
**eFigure 8.** Histograms of the Differences in Three Census Tract Variables Between 2011 and 2017
**eReferences.**

### eMethods. Data and Analysis Methods

#### 1. Recoding Los Angeles County Health Survey (LACHS) measures to create variables used in analysis

The primary LACHS outcome variables used in this study were fast food (FF) intake frequency, obesity, and diabetes, and sociodemographic variables, and they were transformed and/or re-coded from the original measures for ease of interpretability. In addition to the text below, all measures coded by LACHS included categories for "do not know" or "refused". These responses were categorized as "unknown" (**eTable 6**) and were not included in the analysis.

***FF intake frequency***: Respondents were asked, "How often do you eat any food, including meals and snacks, from a fast-food restaurant like McDonald's, Taco Bell, Kentucky Fried Chicken, or another similar type of place?" (1=4 or more times a week, 2=1-3 times/week, 3=less than once a week but more than once a month, 4=less than once a month, 5=never). Because of a small number of respondents in the "4 or more times a week" category (N=300, 3.7%), we re-coded this variable into the 4-category "FF intake frequency" variable analyzed in this study: never, infrequent (< once per month), moderate (≥ once per month to < once per week), and frequent (≥ once per week).

***Obesity***: Respondents reported their current height (inches) and weight (pounds), which was used to calculate Body Mass Index (BMI = $kg/m^2$). A variable for BMI status was coded by the LACHS across 4 categories based on the definitions from the National Heart, Lung, and Blood Institute (NHLBI)[1]: Obese (BMI ≥ 30), Overweight (25 ≤ BMI < 30), Normal Weight (18.5 ≤ BMI < 25), Underweight (BMI < 18.5). We re-coded this into the binary variable for "obesity" used in this study: 1=yes, 2=no.

***Diabetes***: Respondents were asked "Have you ever been told by a doctor or other health professional that you have diabetes or sugar diabetes [IF FEMALE, ADD: other than during pregnancy]?", coded by LACHS and used as the "diabetes" variable in this study: 1=yes, 2=no.

***Sociodemographics***:

(i) **Gender**: Gender was recorded by LACHS as male or female, used as the "gender" variable in this study.

(ii) **Age group**: "What is your age?" (numerical response in years) was recorded by LACHS with the categories: 18-24; 25-29; 30-39; 40-49; 50-59; 60-64; and 65 or over, used as the "age group" variable in this study.

(iii) **Race/ethnicity**: Race was measured by asking participants, "What is your race?" (White; Black/African American; Asian; Pacific Islander; American Indian/Alaskan Native; Hispanic/Latino; Other; Do not know; Refused). Hispanic origin was measured by asking participants, "Are you of Latino or Hispanic origin?" (yes, no). These two variables were coded by LACHS into a combination variable with 7 categories: Hispanic/Latino; White; Asian; Black/African American; Native Hawaiian or Pacific Islander; American Indian/Alaskan Native; Other, based on the following rules:

- If Hispanic/Latino mentioned at all, Hispanic/Latino was assigned.
- Else if Black/African American mentioned at all, African American was assigned.
- Else if Pacific Islander mentioned at all, Native Hawaiian or Other Pacific Islander (NHOPI) was assigned.
- Else if Asian mentioned at all, Asian was assigned.
- Else if White mentioned only, White was assigned.
- Else if American Indian/Alaska Native mentioned only, American Indian/Alaska Native (AI/AN) was assigned
- The remaining was assigned as Other.

We re-coded the combination variable into the 5-category "race/ethnicity" variable used in this study: Hispanic/Latino; White; African American; Asian; and Multiracial/Other, which includes Pacific Islander, American Indian/Alaskan Native, do not know, and refused.

(iv) **Household income level**: Participants were asked for their annual household income. Responses were recoded by LACHS into a 4-category variable for household income level relative to the Federal Poverty Level (FPL): 0-99% FPL; 100-199% FPL; 200%-299% FPL; 300% or above FPL. We recoded this 4-category variable into the binary variable for "household income level" used in this study: low-income = <200% of the FPL, high-income = ≥200% FPL.

(v) **Education level**: "What is the highest level of school you have completed or the highest degree you have received?" was coded by LACHS into the 4-category variable: 1='Less than high school'; 2='High school'; 3='Some college or trade school'; 4='College or post graduate degree'.

***Census tract***: Respondents reported their home address or nearest cross-streets to their home. Addresses were geocoded with street information to census tracts. When this information was not provided, the census tract was coded as missing data.

#### 2. Mobility data processing

##### 2.1 Data collection

Geolocation (i.e., mobility) data were collected by Cuebiq, a location intelligence and measurement company that supplies a patented software development kit (SDK) to mobile app developers, providing a privacy compliant path for anonymous users to opt-in to share location data. Prior studies have used this particular individual-level data resource to study inequality in use of urban spaces[2] and to model the impact of non-pharmaceutical interventions during the COVID-19 epidemic[3].

The data are collected from anonymized users who have opted in to provide access to their GPS location to specific applications (apps) using Location Based Services (LBS). Permitting users have selected phone settings to allow LBS to be activated when specific apps are in use. Cuebiq collaborates with multiple smartphone app providers to access location information anonymously through a General Data Protection Regulation (GDPR) and California Consumer Privacy Act (CCPA) compliant framework. Devices across all major operational systems (e.g., iOS, Android, Windows), are included, although Android devices make up the majority of the data collected.

Data was shared in 2017 under a strict contract with Cuebiq through their Data for Good program[4], which provides access to anonymized and privacy-protected mobility data for academic research and humanitarian initiatives only. All researchers were contractually obligated never to attempt to de-identify data, single out identifiable individuals, or link these data to third-party data about an individual. All study protocols were approved by the Institutional Review Boards (IRBs) of the Los Angeles County (LAC) Department of Public Health, the University of Southern California, and Massachusetts Institute of Technology.

##### 2.2 Data accuracy

Each device frequently broadcasts its location to a central server by sending its latitude, longitude, device ID, and the exact date and time of the event, which together represents a time-location *ping*. The ping data from Cuebiq also come with an estimate of the *horizontal positioning accuracy*, or accuracy, of each ping. For data collected from Android devices, which are the primary source of location data for this dataset, accuracy is defined as the radius of 68% confidence for the location, measured in meters (m). This means that if a circle is drawn centered at the latitude and longitude specified by the ping, with radius equal to the accuracy, there is a 68% probability that the true location is inside the circle[5]. The definition of horizontal positioning accuracy is similar for data collected from iOS (Apple) devices[6].

**eFigure 1** presents a frequency distribution of the accuracy of all pings generated by a random sample of 2,000 (0.8%) of smartphone users in our dataset with accuracy levels ranging from 0 to 200m. The median accuracy of this distribution is 21m. An accuracy of 0 means that the accuracy level of the ping is < 1m. Pings with accuracy > 200m were not included in analysis for this study.

More generally, several studies have evaluated the accuracy of GPS data collection via crowdsourced Android and Apple devices. Accuracy has been found to be affected by various conditions including whether the smartphone is using GPS only or GPS combined with WiFi network, and the level of activity on that network; is within specific indoor locations; or outdoor locations (e.g., under leafy coverage, or in the presence of multiple multi-story buildings)[7,8]. A 2019 study found the average accuracy of location measurements in urban environments across two times of day and two times of year using an iPhone 6 to be between 7 - 13m; this is consistent with the accuracy range observed of recreation-grade GPS receivers (e.g., Garmin)[7]. While iPhone 6 was discontinued in 2016, it was still a prevalent model in use during the 2016-2017 data collection period. More recent iPhone models may have more highly resolved accuracy.

##### 2.3 Detecting stays

When a user spends significant time at a single location, measurement uncertainty will cause a number of pings to be scattered around the actual location. To map these events to a single stay with an accurate time and location, we use the Infostop algorithm[9]. Extracting stays compresses ping trajectories into a series of stay points coded with the attributes latitude, longitude, start time, and end time. To extract the locations of stays, the algorithm clusters consecutive events together if the maximum distance from their centroid, computed as the median of the pings' latitudes and longitudes, is less than some roaming distance, $d^{roam}$. The first and last ping mark the start and end time of the stay. At least two subsequent events need to be observed within $d^{roam}$ to be considered a stay. To better estimate the location of places that are visited frequently by the same user, the algorithm also checks whether different clusters appear within $d^{roam}$ of each other and assigns a single consistent location to all connected clusters by recomputing their centroid. We use $d^{roam}$ = 50m, and set the minimum duration of a stay to be 5 minutes.

**eTable 1** provides statistics on the distribution of individual stays in the mobility data, based on the analytic sample of n=234,995 users observed between October 1, 2016, and March 31, 2017, 182 days. Across our sample we find a total of 63,299,255 stays at locations within LAC, with a median (interquartile range [IQR]) number of stays per user of 172 (IQR, 93, 320). These stays were collected across a total of 16,009,417 observation days, with a median of 57 (IQR, 34, 90) days of observation for each user.

##### 2.4 Attributing stays to Points of Interest (POI)

A key feature of our analysis is how we model the attribution of stays detected in the mobility data to specific Points of Interest (POI), some of which are food outlets. Like stays, each POI is represented by a single point in space. To attribute a stay to a POI, we attribute each stay to the closest POI in our dataset (discussed below), calculated from the centroid of the user's stay to the centroid of the spatial polygon of the POI. This approach has been demonstrated to be accurate for inferring visited POI from passively collected smartphone mobility data[10]. To avoid attributing a stay to a distant POI, we choose only the closest POI within a radius of a thresholding maximum distance, $d^{max}$, which we set at 200m. If a stay is further than $d^{max}$ from any venue, the stay is discarded. Although we set $d^{max}$= 200m, the median distance of stays attributed to POI, $d^{stay}$, is 29.8 m (IQR, 14.7 - 55.4), and the median distance of a stay attributed to food outlet POIs (defined below) specifically is 27.2 m (IQR, 12.5 - 67.0).

We have tested the robustness of our results to this approach for attributing stays to POI by exploring values of $d^{max}$ smaller than 200m: $d^{max}$ = 20, 50, 100, and 200m. Since the average accuracy of this data may be as high as 13m, we do not explore values of $d^{max}$ below 20m. **eTable 4** presents statistics on the total number of stays and the median (IQR) distance $d^{stay}$ between a stay and attributed POI at each value of $d^{max}$. Applying a $d^{max}$ of 20m or 50m is very restrictive, as evidenced by the low number of stays attributed and median values of $d^{stay}$ under these conditions.

We perform robustness tests at two levels of analysis. First, we evaluate the correlation between the values of each FF outlet visit variable at each value of $d^{max}$. Specifically, we test the Pearson correlation coefficient between the values of the FF visits/time variable in each of the LAC neighborhoods calculated at $d^{max}$=200 and all other values of $d^{max}$; we repeat this analysis for the FF visits/food variable. The correlations between the two variables are very high for all values of $d^{max}$. For FF visits/time, we find $\rho[d^{max}=200m, d^{max}=100m] = 0.98$, $\rho[d^{max}=200m, d^{max}=50m] = 0.96$, and $\rho[d^{max}=200m, d^{max}=20m]=0.91$; for FF visits/food we find $\rho[d^{max}=200m, d^{max}=100m] = 0.99$, $\rho[d^{max}=200m, d^{max}=50m] = 0.97$, and $\rho[d^{max}=200m, d^{max}=20m] = 0.93$ (**eFigure 3**). These results show that the FF outlet visit variables are largely independent of the details of our approach for attributing stays to POI.

We also test the robustness of the results of the association between FF outlet visits and FF intake frequency to the value of $d^{max}$ (**eTable 5**). Specifically, we (i) calculated values of the FF outlet visit variables at each $d^{max}$ across the LAC neighborhoods, (ii) linked these to the analytic sample of LACHS users based on home census tract of residence, and (iii) fit regression models of the association between FF intake frequency and the FF visit variables calculated at each value of $d^{max}$. Comparing results across values of $d^{max}$, we find very slight fluctuations in estimated OR from regression analysis (**eTable 5**). These results indicate that our main findings are largely independent of and robust to the details of our approach for attributing stays to POI.

##### 2.5 Identifying food and fast food outlets

We obtain the location of POI and food outlets in LAC using a large places database obtained from the technology company Foursquare via their Public Search API[11] in 2017 and according to their terms and conditions of use. Foursquare, now called Foursquare City Guide, popularized the concept of real-time location-sharing and checking-in[12]. The data is built from a combination of crowd-sourced user activity and the aggregation of data from additional sources[13]. A 2018 study comparing the Foursquare POI database with other public POI databases from mapping and social media platforms (Facebook, Foursquare, Google, Instagram, OSM, Twitter, and Yelp) established that while none of these databases is complete, Foursquare's data quality, as measured by number of POI, number of categories included, and positioning accuracy, was among the best[14].

The POI database we downloaded in 2017 provides the names and geolocation of n=239,509 POI in LAC. These POI came classified by Foursquare across ten primary categories, *pcat* (e.g., *pcat* = Food; Shop & Service; Nightlife Spot; College & University; etc.) and 665 subcategories, *cat* (e.g., for *pcat*=Food, *cat* = Fast Food; American; Burger; Vietnamese; etc.)[1].

We took several approaches to modify and recode this existing taxonomy to define the categories of *food outlets* and *FF outlets* analyzed in this study. We define a *food outlet* as any location where food might be sold. We start by accepting all POI falling under Foursquare's existing primary category of *pcat=Food*. We then combed through all subcategories under a primary category not equal to *Food* (e.g., *pcat = Nightlife Spot*) to identify additional locations where food might be sold, which we re-coded as *food outlets*. **eTable 2** shows all the non-food primary category and subcategory combinations coded as *food outlets*.

To define the *FF outlet* category, we start by accepting all POI within the primary category *cat=Food* and subcategory *cat=Fast Food*. We enrich this Foursquare-defined list by performing a search of known chain FF outlets validated in previous nutritional health research as representing limited-service restaurants serving menus of predominantly ultra-processed and/or low-nutrient, energy dense foods[15,16] (**eTable 3**). The search was performed by matching substrings from the list in **eTable 3** with the names of POI in the Foursquare database. After re-coding, we find a total of 53,588 *food outlets* and 4,151 *FF outlets* in LAC out of the n=239,509 POI in the Foursquare database.

In comparison, the LAC Restaurant and Market Inventory[17], which comprises Environmental Health permitted restaurants and markets in LAC that are inspected by the LAC Department of Public Health, contains a total of 40,600 restaurants and markets, approximately 13,000 fewer than in our Foursquare database.

##### 2.6. Discussion of mobility data processing

We have taken steps to (i) validate the measurement accuracy of the mobility data (eMethods Sections 2.2-2.3), (ii) validate the robustness of findings to our methods for attributing geolocations to particular POI (eMethods Section 2.4), and (iii) define, detect, and appropriately label food and FF outlets (eMethods Section 2.5). However, there may be limitations to our ability to detect visits to certain food outlets, such as those in particularly dense urban areas or multi-story or multi-purpose buildings (e.g., malls) where FF outlets may be concentrated. Additionally, because we only detect visits greater than five minutes in duration, we may miss brief FF outlet visits (e.g., drive throughs).

The Foursquare POI database we use has limitations, including coverage of food outlets that are infrequently visited and spatially dynamic outlets like food trucks. Yet, it is well-established that all food environment databases have limitations[18], and we have demonstrated advantages of this database including that it is more comprehensive than the LACDPH-maintained inventory of permitted food-selling establishments[17], and referencing previous work demonstrating that its quality is among the best of any publicly available POI database in 2018, close to the time of our data collection in 2017[14].

#### 3. Mobility data representativeness

##### 3.1 Neighborhood-level aggregation of mobility-derived variables and representativeness

Mobility measures were aggregated and averaged across users within spatial areas. Aggregation to an area-level was necessary because privacy protections set out in the MIT and USC IRB protocols did not allow reporting on the behavior of individual mobility users. We explored aggregating the users over two existing administrative spatial boundary divisions: the U.S. Census Bureau U.S.-wide census tract level, with 2,346 units within LAC; and the neighborhood level, specific to LAC and with 272 neighborhood units (described in eMethods Section 3.1.1). We investigated the sufficiency of the mobility user sample size within boundaries at each level (eMethods Section 3.1.2), and whether broad population representativeness is achieved at each level (eMethods Section 3.1.3). We tested these two questions and based on the results, decided to aggregate smartphone users at the neighborhood level. As a final step, we used post-stratification weighting to adjust the sampling of FF outlet visit variables within neighborhoods to appropriately represent the census tracts composing each neighborhood (described in eMethods Section 3.1.4).

[1] For a list of all venue categories see https://developer.foursquare.com/docs/build-with-foursquare/categories/

##### 3.1.1 Background on LAC neighborhoods

The LAC neighborhoods were designed as part of a community-sourced project to map LAC communities, led by the Los Angeles Times (LA Times) Newspaper Datadesk[19]. They were crafted with the goal of representing communities across LAC with more similar groups of people. Neighborhoods are classified into three "types": *segment-of-a-city*, *standalone-city*, and *unincorporated-area*. To define neighborhoods across the county, the LA Times started with the U.S. Census Bureau's boundaries of 88 cities and 43 census-designated places and worked with its readership to determine which cities to keep as a *standalone-city*, such as Santa Monica; and which to subdivide into multiple *segment-of-a-city*. Multiple cities were divided into *segment-of-a-city*, with the most notable example being Los Angeles City (population approximately 4 million, 40% of the LAC population), which is divided into 114 *segment-of-a-city*. The *segment-of-a-city* were created by joining neighboring census tracts within the city boundaries through an iterative mapping process with the LA Times' readership. Census-designated places were either kept as independent areas, categorized as an *unincorporated-area*; or, if they closely adjoin or are entirely within a city boundary, were combined with those cities. The median (IQR, range) number of residents in each neighborhood is 27,499 (IQR, 12,961-53,124; range, 58-471,568). A map of the resulting neighborhood boundaries is provided in **eFigure 2**. The median (IQR) land area of the neighborhoods in square miles ($mi^2$) is 3.63 (IQR, 1.84-8.78) $mi^2$, and the range is 0.26-442.00 $mi^2$.

##### 3.1.2 Mobility user sample size sufficiency

We investigated whether the sample size of mobility users within each boundary designation was large enough to achieve stable estimates of mobility measures. This was tested by comparing the distributions of the size of the mobility user sample in each census tract with that within each neighborhood level (**eFigure 4**).

Of the 2,346 census tracts in LAC, 2,266 had residing smartphone users in our sample (based on estimated home address) and did not have a rural designation[20]. The median (IQR, range) number of users per census tract is 71 (IQR, 52-95; range, 12-323); 80% of census tracts have fewer than 100 users. Of the 272 neighborhoods in LAC, 247 had residing smartphone users and were not composed of a majority of rural census tracts[20]. The median (IQR, range) number of users per neighborhood is 468 (IQR, 243-914; range, 14-7,326); 7% of neighborhoods have fewer than 100 users.

We concluded that the small sample of smartphone users within each census tract jeopardizes the ability to reach a stable estimate of the FF outlet visit variables within these areas, especially when considering that only a subset of users within each area will have logged any observations of FF outlet visit behavior.

##### 3.1.3 Population representativeness

We investigated whether broad geographic representation of the underlying population size at the census tract vs. neighborhood level was achieved by the mobility user sample. This was tested by comparing the Pearson correlation between the number of users in the mobility dataset and the underlying population reported in the 2017 American Community Survey (ACS 2017)[21] at the census tract and the neighborhood levels. **eFigure 5a-b** are scatter plots between ACS population and user population at the census tract and neighborhood levels, respectively, also showing the Pearson correlation. We find that the ACS population and number of mobility users are correlated at the census tract level with Pearson $\rho = 0.66$ ($P<.001$), and at the neighborhood level with Pearson $\rho = 0.97$ ($P<.001$). These results suggest that the mobility data sample is highly representative of the overall population size across the neighborhoods, and less so at the census tract level.

Because of the small sample size of mobility users within the majority of census tracts, and the fact that broad geographic representation of the mobility user sample was achieved over the neighborhoods, we decided to aggregate smartphone users at the neighborhood level.

##### 3.1.4 Post-stratification sampling to represent neighborhood-level populations

We furthermore address the representativeness of the data using post-stratification sampling[22]. Post-stratification is a sampling tool that uses weighting to adjust raw observational data to meet known population or demographic distributions, and is commonly used by computational social science researchers in application to various datasets with broad population coverage, including mobile phone data[2,23]. The essence of the technique is to divide the observed sample into post-strata, and then to compute a weight for each post-strata or case within a post-strata before bringing the post-strata back together to compute statistics for the overall sample.

We performed post-stratification sampling to appropriately represent the true population size of the census tracts that compose each neighborhood. We estimate the arithmetic mean of the FF visit variables within each census tract, and then find the overall mean for the neighborhood by weighting according to the ratio of the census tract population size to the neighborhood population size, as determined by the ACS.

Let $CT_j$ be a census tract, $\overline{FF_i^{CT_j}}$ be the mean of a FF outlet visit variable calculated over $CT_i$, and $NB_k$ be a neighborhood. Then we can find the post-stratified mean of the FF outlet visit variable within $NB_k$, $\overline{FF_i^{NB_k}}$, as

$$\overline{FF_i^{NB_k}} = \sum_{CT_j \in NB_k} \overline{FF_i^{CT_j}} \cdot \frac{\omega^{CT_j}}{\omega^{NB_k}},$$

where $\omega$ represents population size. **eFigures 6a-b** show how FF visits/time and FF visits/food calculated at the neighborhood level change when the post-stratified (i.e., weighted) values are used instead of the unweighted values. The Pearson correlation coefficient between the weighted and unweighted values is $\rho \simeq 0.99$ for each FF outlet variable. This high correlation between unweighted and weighted FF outlet visit variables means that the unweighted data was almost fully representative of the population of each census tract composing each neighborhood. Post-stratifying the sample may alleviate any remaining bias of the data in not appropriately representing the size of the composing census tracts within each neighborhood.

##### 3.2 Representativeness of the mobility data at the level of individual POI

In previous published work on this mobility dataset, we investigated whether our method for detecting when a user spends time at a particular POI is accurate[2]. We devised a test of the ability of our data and attribution approaches to detect visits to an individual POI in a way that is representative of the overall population. The test compares the official attendance counts at games of the major professional sports leagues to the estimates of attendance using our data. Our own estimates were based on (i) the number of individuals that have a stay within the perimeter of the large stadium polygon perimeter between 3 hours before starting time to 3 hours after the game's completion, and (ii) the representativeness of the mobility data to the overall population size. This test was conducted across games of the National Football League (NFL), National Basketball Association (NBA), and National Hockey League (NHL) in LAC, as well as other cities in the U.S. We found that estimates of attendance computed using our data were extremely close to official attendance counts, indicating that this mobility data achieves good representation of visits to large POI.

##### 3.3 Demographic representativeness

In previous published work on this mobility dataset, we investigated whether the user data is representative of the real population distribution of income[2]. We first estimated the income level for each smartphone user, and compared the distribution of estimated income levels across our sample of smartphone users to the distribution of income at the census block group (CBG) level, where official CBG estimates came from the ACS. This test was conducted within LAC and other cities in the U.S. We found that our sample of users has an average income that is 8.6% higher than the census data. This indicates that our sample of mobility users has low bias towards income classes after estimating an income level for each user.

We have not repeated this test with other demographic variables such as race and ethnicity or gender for reasons of user privacy and protection; whether these types of demographic variables should be imputed from digital trace data is an active topic of debate and discussion in the field of Computational Social Science[24,25].

#### 4. Constructing mobility variables

##### 4.1 Temporal frequency of FF outlet visits (FF visits/time)

To maximize the amount of user activity that can be analyzed while accounting for gaps in observation over time, we define a unique observation set for each user, and define the FF outlet visit variables relative to this observation set. We separate all days into three time periods: morning (12:00am -10:59am), midday (11:00am - 3:59pm), and evening (4:00pm - 11:59pm), for a set of $P$ possible daily periods; if there are $d$ days in our observation set, we now have $|P| \leq 3d$. For each user $i$, we identify the subset of $P$ in which $i$ had at least one stay, denoting this $P_i^{stay}$. We then identify the subset of $P_i^{stay}$ in which user $i$ had at least one visit at a FF outlet, denoting this as $P_i^{FF}$. We find the temporal frequency of visits to FF outlets for user $i$, i.e. the FF visits/time variable specific to user $i$,

$FF_i^{time}$, as $FF_i^{time} = \frac{P_i^{FF}}{P_i^{stay}}$. The theoretical range for $FF_i^{time}$ is [0,1]; $FF_i^{time} = 1$ if user $i$ is observed to visit FF in every observed time period.

We obtain the variable FF visits/time at the level of spatial area $A$, $FF_A^{time}$, as the mean of $FF_i^{time}$ over all $N$ users residing within $A$,

$$FF_A^{time} = \frac{1}{N}\sum_{i \in A} FF_i^{time}.$$

##### 4.2 Relative frequency of FF outlet visits (FF visits/food)

To define the relative frequency of FF outlet visits variable, FF visits/food, we consider all observations of visits to food and FF outlets, and do not use the possible daily periods $P$ designation. For user $i$, we define the total number of visits to food outlets as $V_i^{food}$. We define the total number of visits to FF outlets as $V_i^{ff}$. We find the relative frequency of FF outlet visits for user $i$, i.e., the FF visits/time variable specific to user $i$, $FF_i^{food}$, as $FF_i^{food} = \frac{V_i^{ff}}{V_i^{food}}$. The theoretical range for $FF_i^{food}$ is [0,1], and is equal to 1 if all of user $i$'s food visits are to FF outlets.

We obtain the variable FF visits/food at the level of spatial area $A$, $FF_A^{food}$, as the mean of $FF_i^{food}$ over all $N$ users residing within $A$,

$$FF_A^{food} = \frac{1}{N}\sum_{i \in A} FF_i^{time}.$$

##### 4.3 Average number of trips per day (trips/day)

Let $T_i$ be the total number of trips that user $i$ takes during the $d$ days of observation. We find the average number of trips per day across all users $i$ within area $A$, $\overline{T_A}$, as

$$\overline{T_A} = \frac{1}{N}\sum_{i \in A}\frac{T_i}{d}.$$

##### 4.4 Scaling of mobility variables

All continuous variables from the mobility data are rescaled between [0,10] by min-max scaling using the formula, generic for variable X(x) that is scaled into a variable $X'(x')$,

$$X'(x') = 10\,\frac{x - min(X)}{max(X) - min(X)}.$$

A one-unit increase in the scaled variable $X'(x')$represents a 10% increase in the unscaled observed variable $X(x)$.

#### 5. Equations for regression models

logit*(FF INTAKE FREQUENCY)* = $\beta_0 + \beta_1 X_i + \beta_2 AGE + \beta_3 GENDER + \beta_4 RACE + \beta_5 INCOME + \beta_6 EDUCATION$
logit*(OBESE)* = $\beta_0 + \beta_1 X_i + \beta_2 AGE + \beta_3 GENDER + \beta_4 RACE + \beta_5 INCOME + \beta_6 EDUCATION$
logit*(DIABETES)* = $\beta_0 + \beta_1 X_i + \beta_2 AGE + \beta_3 GENDER + \beta_4 RACE + \beta_5 INCOME + \beta_6 EDUCATION$

where $X_i \in$ {FF visits/time, FF visits/food, FF intake frequency}

#### 6. Sensitivity analyses

##### 6.1 Adjusting for General Mobility Behavior

To examine whether the observed relationships between visits to FF outlets and FF intake and diet-related disease are uniquely due to FF outlet visits, and not general mobility behavior irrespective of FF visits, we test models that control for an indicator of general mobility, the average number of trips per person per day (trips/day).

After adjusting for general mobility behavior in models predicting both obesity and diabetes, there is almost no change in the effect sizes for FF visits/time and FF visits/food (**eTable 8**).

##### 6.2 Evaluating model sensitivity to changes in demographic distribution of underlying LAC population between 2011 and 2017

###### 6.2.1 Methods

Relating measures of mobility to food outlets in 2016/17 with responses from a 2011 health survey represents a possible source of error in our approach. We conducted sensitivity tests to examine the compatibility between the two datasets and whether any incompatibility due to differences in neighborhood populations between LACHS data collection in 2011 and mobility data collection in 2016-17 could have contributed error to model results. The tests firstly investigated how much the demographic distribution of the U.S. Census population[21] changed over this time period, and then estimated the effect of this changing distribution on model results. We focused on variables representing the percentage of the population in the census tract: (i) living above 200% of the FPL, (ii) that is Hispanic, and (iii) that is African-American, all of which are strong predictors of FF intake frequency. The percentage of the population living above 200% of the FPL was used as a measure of income because it accounts for inflation and corresponds to the income measure available for LACHS study respondents.

We measured the distribution of differences in each of these three percentage-based measures between 2011 and 2017. We then identified LAC census tracts that were considered 'outliers', i.e., the census tracts that demonstrated the greatest amount of change according to two methods: (1) the Tukey approach for outlier identification, which identifies outliers as values more than 1.5 times the interquartile range from each of the quartiles (method 1); and (2) by removing the distribution above and below two standard deviations of the mean (method 2). For each outlier identification method, we: (i) removed the union of outlier census tracts across the three measures and the LACHS respondents they were linked to; (ii) re-estimated the regression models presented in the main text after removing the linked LACHS study participants from these census tracts; and (iii) computed *P* values to test whether the differences between odds ratios from regression models fit to the full analytic sample and the sample after subtracting out outlier census tracts according to the two methods were significant, using a $\chi^2$ test.

###### 6.2.2 Results

Between 2011 and 2017, the majority of census tracts did not change in their distribution of demographic features. **eFigure 8** shows the distributions of the differences for each demographic variable. Each distribution resembles a normal distribution and is mean-centered at approximately 0. Ninety-five percent of census tracts demonstrated less than a 16% change in the percentage of the population above 200% of the FPL; a 10% change of the population that is African-American; and a 15% change of the population that is Hispanic.

**eTable 9** shows, for each outlier detection method, the number of census tracts removed and the number of linked LACHS respondents removed from the population sample. Approximately 12.4% of LACHS respondents (accounting for up to 12% of census tracts) were removed as outliers, i.e., as demonstrating the greatest amount of change in the three demographic variables considered.

Regression analyses were rerun for six models representing the primary results analyzed in this study after removing LACHS respondents in outlier census tracts. These models examined the association between each of the two FF outlet visit measures observed from the mobility data, FF visits/time and FF visits/food, and each of the three outcome measures analyzed, FF intake frequency, obesity, and diabetes. We adjusted for demographics in the obese and diabetes outcome models. Regression results were compared between results using the original analytic sample and the analytic sample after removing outliers by the two outlier-detection methods.

**eTable 10** presents unadjusted OR from multinomial regression models of self-reported FF intake fit to the full analytic sample, compared with models fit to the analytic sample subtracting out outlier census tracts according to method 1 and 2. The *P* values for differences between ORs obtained on the full sample and the sample subtracting out outlier census tracts are presented in **eTable 11**. For all OR, using both methods for identifying outlier census tracts, differences in ORs from regression models fit to the full analytic sample vs. the model without outliers were not significant.

**eTable 12** and **eTable 13** present adjusted OR from binary regression models of obesity and diabetes, respectively, fit to the full analytic sample, compared with results from regression models fit to the analytic sample subtracting out outlier census tracts according to method 1 and 2. The *P* values for differences between ORs obtained on the full sample and the sample subtracting out outlier census tracts are presented in **eTable 14**. For all OR, both obesity and diabetes outcomes, and both methods for identifying outlier census tracts, differences in OR from regression models fit to the full analytic sample were not significant.

**eTable 1. Statistics of the Distribution of Individual Stays in the Mobility Data Across the Analytic Sample of Smartphone Users***

| Quantile | Number of stays | Number of days with stays |
|---|---|---|
| **Min** | 1 | 1 |
| **25%** | 93 | 34 |
| **Median** | 172 | 57 |
| **75%** | 320 | 90 |
| **Max** | 6,706 | 182 |
| **Total** | 63,299,255 | 16,009,417 |

* Across the analytic sample of n=243,644 users in the mobility data observed between October 1, 2016, and March 31, 2017, 182 days.

**eTable 2. Non-Food Primary Category and Subcategory Combinations Coded as *food outlets* in the Foursqaure dataset**

| **Primary Category, *pcat*** | **Subcategory, *cat*** |
|---|---|
| Nightlife Spot | American, Asian, Bar, BBQ, Beach Bar, Beer Bar, Beer Garden, Brewery, Burgers, Café, Cocktail, French, Hookah Bar, Hotel Bar, Italian, Japanese, Korean, Lounge, Mediterranean, New American, Pizza, Sake Bar, Seafood, Speakeasy, Sports Bar, Sushi, Tiki Bar, Whisky Bar, Wine Bar, Wine Shop, Winery, Convenience Store, Dive Bar, Gastropub, Liquor Store, Mexican, Pub, Restaurant, Steakhouse, Wings |
| College & University | Café, Coffee Shop |
| Arts & Entertainment | American, Café, Cocktail, Dive Bar, Piano Bar, Pub, Restaurant, Speakeasy, Bar, Lounge, Pizza |
| Outdoors & Recreation | Farmer's Market, American, Speakeasy |
| Professional & Other Places | American, Cafeteria, Coffee Shop, Wine Bar, Winery, Corporate Cafeteria, Corporate Coffee Shop |
| Shop & Service | American, Beer Store, Candy Store, Cheese Shop, Chocolate Shop, Convenience Store, Deli / Bodega, Desserts, Discount Store, Farmer's Market, Fish Market, Food & Drink, Fruit & Vegetable Store, Gourmet, Grocery Store, Health Food Store, Juice Bar, Mexican, Organic Grocery, Restaurant, Sandwiches, Smoothie Shop, Snacks, Street Food Gathering, Supermarket, Warehouse Store, Wine Bar, Wine Shop, Bakery, Butcher, Café, Herbs & Spices Store, Liquor Store |
| Travel & Transport | Food Truck, Hotel Bar |

#### eTable 3. List of Fast Food Outlet Names*

| | | | | |
|---|---|---|---|---|
| Deangelo Pizza | Angelo Sandwich Shop | Schlotzky | Hardee | Burger King |
| Papa Gino | Jack In The Box | Dairy Queen | KFC | El Pollo Loco |
| Dominos | Bob Big Boy | Sonic | Kentucky Fried Chicken | Carl's Jr. / Green Burrito |
| Godfather Pizza | In-N-Out Burger | Blimpie Subs And Salads | KFC/Taco Bell | Carl's Jr./Green Burrito |
| Papa John | Five Guys | Waffle House | Wienerschnitzel | Carl's Jr. |
| Pizza Hut | Bob's Big Boy | Chick-Fil-A | Fuddruckers | Green Burrito |
| Pizza Inn | Checkers Drive-In Restaurant | Whataburger | Long John Silver | Baja Fresh |
| Shakey Pizza | Bojangles | Del Taco | Arby | Waba Grill |
| Papa Murphy | Denny | In N Out Burger | Harvey | Wendy's |
| Little Caesars | Steak N Shake | Krystal | Subway | Arby's |
| Pizza Delight | Togo | Buffalo Wild Wings | Orange Julius | Bob's Big Boy |
| Pizza Pizza | Popeye | Cici Pizza | Taco Bell | Rally's |
| Chuck E Cheese Pizza | Church Chicken | Frisch Big Boy | Taco John | Yoshinoya |
| Mr Gatti Pizza | White Castle | Hungry Howie | Wendy | Weinerscnitzel |
| Round Table Pizza | Carl Jr | Rally Hamburgers | Mc Donald | Sonic Drive-In |
| TGI Friday's | Quizno | Rally's | Mcdonald's | |

* Validated in previous nutritional health research as representing limited-service restaurants serving menus of predominantly ultra-processed and/or low-nutrient, energy dense foods[15,16], and used to enrich Foursquare's existing food outlet categorization.

#### eTable 4. Statistics of Stays at Each Value of Thresholding Maximum Distance

| | Stays at any POI | | | | Stays at food outlet POI | | | |
|---|---|---|---|---|---|---|---|---|
| | Threshold for maximum distance between smartphone user and Point of Interest, $d^{max}$ (meters) | | | | | | | |
| **Statistic** | **20** | **50** | **100** | **200*** | **20** | **50** | **100** | **200*** |
| Total number of stays | 17,484,338 | 35,367,227 | 49,955,726 | 63,299,255 | 5,781,210 | 9,803,413 | 12,296,092 | 14,498,850 |
| Distance from stay to POI, $d^{stay}$ median (IQR) | 6.3 (4.2, 8.2) | 11.1 (6.9, 15.3) | 20.2 (11.2, 32.2) | 29.8 (14.7, 55.4) | 10.3 (6.3, 14.7) | 16.6 (9.1, 27.8) | 21.6 (10.8, 43.0) | 27.2 (12.5, 67.0) |

Abbreviations: IQR, interquartile range.
* Results in the main paper were calculated at $d^{max}$ = *200m*.

#### eTable 5. Unadjusted Odds Ratios of Self-Reported Fast Food Intake in Robustness Analyses of the Attribution of Stays Detected in Mobility Data to Points of Interest[a]

| | **FF intake frequency, unadjusted OR (95% CI)** | | | | | | | | | | | |
|---|---|---|---|---|---|---|---|---|---|---|---|---|
| | **Infrequent** | | | | **Moderate** | | | | **Frequent** | | | |
| | **Threshold for maximum distance between smartphone user and Point of Interest, $d^{max}$ (meters)** | | | | | | | | | | | |
| **Model[b]** | **20** | **50** | **100** | **200[c]** | **20** | **50** | **100** | **200[c]** | **20** | **50** | **100** | **200[c]** |
| FF visits/time | 1.12 (1.06, 1.18) | 1.13 (1.06, 1.19) | 1.13 (1.06, 1.19) | 1.13 (1.06, 1.20) | 1.25 (1.19, 1.31) | 1.26 (1.19, 1.32) | 1.25 (1.19, 1.32) | 1.26 (1.19, 1.33) | 1.33 (1.27, 1.39) | 1.34 (1.28, 1.41) | 1.33 (1.27, 1.40) | 1.35 (1.28, 1.42) |
| FF visits/food | 1.13 (1.07, 1.19) | 1.12 (1.06, 1.17) | 1.12 (1.06, 1.17) | 1.12 (1.06, 1.17) | 1.25 (1.19, 1.31) | 1.23 (1.18, 1.29) | 1.23 (1.17, 1.28) | 1.22 (1.16, 1.27) | 1.33 (1.27, 1.40) | 1.30 (1.25, 1.36) | 1.29 (1.24, 1.35) | 1.28 (1.22, 1.33) |

Abbreviations: OR, odds ratio; FF, fast food.

[a] Multinomial logistic regression models for fast food intake frequency across four frequency categories; reference group: never. Values of FF outlet visit variables calculated at various thresholds for the maximum distance between a smartphone user and Point of Interest, $d^{max}$, used in our model for attributing stays detected in the mobility trajectory data to a Point of Interest. We investigate values of $d^{max}$ = *20m, 50m, 100m*, and *200m*. Values of the FF outlet visit variables, calculated at each $d^{max}$ and aggregated over each neighborhood, were linked to the analytic sample of LACHS users based on home census tract of residence. Regression models were fit to the linked data at each threshold. $P < .001$ for all estimated odds ratios.

[b] Each model estimated fast food intake frequency using the fast food visit variable listed in this column as the primary independent variable.

[c] Results in the main paper were calculated at $d^{max}$ = *200m*.

**eTable 6. Demographic, Diet, and Disease Characteristics in the Full and Analytic Samples of LACHS Participants and Differences Based on χ2 Test[a]**

| | Participants, No. (%) | | |
|---|---|---|---|
| Characteristic | **Full Sample (n=8036)** | **Analytic Sample (n=5447)** | ***P* value** |
| Age | | | .005 |
| 18-24 | 596 (7.4%) | 467 (8.6%) | |
| 25-29 | 438 (5.5%) | 341 (6.3%) | |
| 30-39 | 1,204 (15.0%) | 878 (16.1%) | |
| 40-49 | 1,596 (19.9%) | 1,063 (19.5%) | |
| 50-59 | 1,674 (20.8%) | 1,118 (20.5%) | |
| 60-64 | 748 (9.3%) | 464 (8.5%) | |
| 65 or over | 1,780 (22.2%) | 1,116 (20.5%) | |
| Gender | | | .044 |
| Female | 4,863 (60.5%) | 3,201 (58.8%) | |
| Male | 3,173 (39.5%) | 2,246 (41.2%) | |
| Race/ethnicity | | | .002 |
| White | 3,414 (43.4%) | 2,257 (41.4%) | |
| Hispanic/Latino | 2,769 (35.2%) | 2,050 (37.6%) | |

| | | | |
|---|---|---|---|
| African American | 784 (10.0%) | 584 (10.7%) | |
| Asian | 728 (9.3%) | 432 (7.9%) | |
| Multiracial/Other | 174 (2.2%) | 124 (2.3%) | |
| Unknown | 167 | 0 | |
| Education | | | .5 |
| Less than high school | 1,385 (17.4%) | 942 (17.3%) | |
| High school | 1,370 (17.2%) | 965 (17.7%) | |
| Some college or trade school | 2,007 (25.2%) | 1,409 (25.9%) | |
| College or post graduate degree | 3,206 (40.2%) | 2,131 (39.1%) | |
| Unknown | 68 | 0 | |
| Income | | | .034 |
| Low | 2,980 (37.1%) | 2,119 (38.9%) | |
| High | 5,056 (62.9%) | 3,328 (61.1%) | |
| Fast food intake frequency | | | .014 |
| Never | 1,552 (19.4%) | 944 (17.3%) | |
| Infrequent | 1,540 (19.3%) | 1,040 (19.1%) | |

| | | | |
|---|---|---|---|
| Moderate | 2,113 (26.9%) | 1,463 (26.9%) | |
| Frequent | 2,794 (34.9%) | 2,000 (36.7%) | |
| Unknown | 37 | 0 | |
| Obesity | | | .072 |
| No | 5,751 (76.6%) | 4,097 (75.2%) | |
| Yes | 1,757 (23.4%) | 1,350 (24.8%) | |
| Unknown | 528 | 0 | |
| Diabetes | | | >.9 |
| No | 7,125 (88.8%) | 4,841 (88.9%) | |
| Yes | 895 (11.2%) | 606 (11.1%) | |
| Unknown | 16 | 0 | |

Abbreviation used: FF (fast food).
[a] $\chi^2$ test for the statistical significance of differences based on non-missing categories.

#### eTable 7. *P* Value for the Difference Between Unadjusted and Adjusted Odds Ratios From Regression Analyses of Self-Reported Fast Food Intake[a]

| Model[b] | *P* value for difference | | |
| --- | --- | --- | --- |
| | Infrequent | Moderate | Frequent |
| FF visits/time | .83 | .44 | .37 |
| FF visits/food | .75 | .37 | .40 |

Abbreviations: OR, odds ratio; FF, fast food.

[a] *P* values are calculated using a $\chi^2$ test to test for the significance of difference between odds ratios from unadjusted and adjusted multinomial regression models for fast food intake frequency across four frequency categories. Reference group: never.

[b] Each model estimated fast food intake frequency using the fast food visit variable listed in this column as the primary independent variable.

#### eTable 8. Logistic Regression Analyses of the Association Between Visits to Fast Food Outlets With Diet-Related Disease Adjusted for General Mobility Behavior[a]

| | | **Obesity outcome** | | **Diabetes outcome** | |
|---|---|---|---|---|---|
| **Model** | **Variable[b]** | **AOR (95% CI) of obesity** | ***P* value** | **AOR (95% CI) of diabetes** | ***P* value** |
| FF visits/time | FF visits/time | 1.16 (1.12 – 1.21) | <.001 | 1.15 (1.09, 1.21) | <.001 |
| FF visits/time and trips/day | FF visits/time | 1.14 (1.10 – 1.19) | <.001 | 1.14 (1.08 – 1.21) | <.001 |
| | Trips/day | 1.11 (1.05 – 1.18) | <.001 | 1.07 (0.99 – 1.15) | .098 |
| FF visits/food | FF visits/food | 1.13 (1.10 – 1.17) | <.001 | 1.11 (1.07 – 1.16) | <.001 |
| FF visits/food and trips/day | FF visits/food | 1.12 (1.09 – 1.16) | <.001 | 1.11 (1.06 – 1.16) | <.001 |
| | Trips/day | 1.13 (1.07 – 1.19) | <.001 | 1.08 (1.00 – 1.17) | .041 |

Abbreviations: AOR, adjusted odds ratio; FF, fast food.
[a] Binary logistic regression models adjusted for demographics: age group, gender, race/ethnicity, educational level, and household income level.
[b] Each model estimated fast food intake frequency using the variable or combination of variables listed in this column as the primary independent variable.

#### eTable 9. Number of Census Tracts and LACHS Respondents Removed After Identifying Outliers in Census Tract Demographic Change Between 2011 and 2017[a]

| Outlier detection method | N (%) census tracts removed | N (%) LACHS respondents removed |
|---|---|---|
| Method 1[b] | 256 (11.0%) | 676 (12.4%) |
| Method 2[c] | 150 (6.7%) | 652 (12.0%) |

[a] Outliers were identified for variables representing the percentage of the population in the census tract: (i) living above 200% of the FPL, (ii) that is Hispanic, and (iii) that is African American. For each outlier identification method, we removed the union of outlier census tracts across the three measures and the LACHS respondents they were linked to and re-computed the sample distribution characteristics.
[b] Outliers are identified by the Tukey method as values more than 1.5 times the interquartile range from each of the quartiles for a variable, i.e. upper outliers are values of the distribution > Q3 + 1.5*IQR and lower outliers are values < Q1 – 1.5*IQR.
[c] Outliers are identified as values of a variable above and below 2 standard deviations of the mean. For each method, we remove from the LACHS sample all respondents living in outlier census tracts. We then identified the union over outlier census tracts across the three measures, as some census tracts overlapped.

#### eTable 10. Unadjusted Odds Ratios From Regression Models of Fast Food Intake From Sensitivity Analyses to Time Change Between 2011-2017[a]

| | FF intake frequency, unadjusted OR (95% CI) | | | | | | | | |
|---|---|---|---|---|---|---|---|---|---|
| | Infrequent | | | Moderate | | | Frequent | | |
| Model[b] | Full sample | Method 1[c] | Method 2[d] | Full sample | Method 1[c] | Method 2[d] | Full sample | Method 1[c] | Method 2[d] |
| FF visits/time | 1.13 (1.06, 1.20) | 1.12 (1.04, 1.19) | 1.11 (1.04, 1.19) | 1.26 (1.19, 1.33) | 1.28 (1.20, 1.36) | 1.26 (1.19, 1.34) | 1.35 (1.28, 1.42) | 1.38 (1.30, 1.46) | 1.35 (1.27, 1.43) |
| FF visits/food | 1.12 (1.06, 1.17) | 1.11 (1.05, 1.17) | 1.11 (1.05, 1.17) | 1.22 (1.16, 1.27) | 1.24 (1.18, 1.30) | 1.22 (1.16, 1.28) | 1.28 (1.22, 1.33) | 1.30 (1.24, 1.36) | 1.28 (1.22, 1.34) |

Abbreviations: OR, odds ratio; FF, fast food.
[a] Multinomial logistic regression models for FF intake frequency across four frequency categories; reference group: never. Sensitivity tests fit regression models to the analytic sample subtracting out respondents living in outlier census tracts demonstrating the largest change in demographic variables between 2011 and 2017 according to methods 1 and 2. Regression model results fit to the full analytic sample (full sample) are provided for comparison. P <.001 for all estimated odds ratios.
[b] Each model estimated FF intake frequency using the FF visit variable listed in this column as the primary independent variable.
[c] Outliers are identified by the Tukey method as values more than 1.5 times the interquartile range from each of the quartiles for a variable, i.e. upper outliers are values of the distribution > Q3 + 1.5*IQR and lower outliers are values < Q1 – 1.5*IQR.
[d] Outliers are identified as values of a variable above and below 2 standard deviations of the mean. For each method, we remove from the LACHS sample all respondents living in outlier census tracts. We then identified the union over outlier census tracts across the three measures, as some census tracts overlapped.

#### eTable 11. *P* Values for Differences Between Odds Ratios From Regression Models of Fast Food Intake From Sensitivity Analyses to Time Change Between 2011-2017[a]

| | **FF intake frequency, *P* value for difference[b]** | | | | | |
|---|---|---|---|---|---|---|
| | **Infrequent** | | **Moderate** | | **Frequent** | |
| **Model[c]** | **Method 1[d]** | **Method 2[e]** | **Method 1[d]** | **Method 2[e]** | **Method 1[d]** | **Method 2[e]** |
| FF visits/time | .88 | .75 | .77 | 1.00 | .70 | 1.00 |
| FF visits/food | .84 | .83 | .70 | 1.00 | .71 | 1.00 |

Abbreviations: FF, fast food.
[a] Multinomial logistic regression models for FF intake frequency across four frequency categories; reference group: never.
[b] *P* values are calculated using a $\chi^2$ test to test for the significance of differences between odds ratios from multinomial regression models fit to the full analytic sample and from regression models fit to the analytic sample subtracting out outlier census tracts demonstrating the largest change in demographic variables between 2011 and 2017 according to methods 1 and 2.
[c] Each model estimated FF intake frequency using the FF visit variable listed in this column as the primary independent variable.
[d] Outliers are identified by the Tukey method as values more than 1.5 times the interquartile range from each of the quartiles for a variable, i.e. upper outliers are values of the distribution > Q3 + 1.5*IQR and lower outliers are values < Q1 – 1.5*IQR.
[e] Outliers are identified as values of a variable above and below 2 standard deviations of the mean. For each method, we remove from the LACHS sample all respondents living in outlier census tracts. We then identified the union over outlier census tracts across the three measures, as some census tracts overlapped.

#### eTable 12. Adjusted Odds Ratios From Regression Models of Obesity From Sensitivity Analyses to Time Change Between 2011-2017[a]

| | Full sample | | Method 1[c] | | Method 2[d] | |
|---|---|---|---|---|---|---|
| **Model[b]** | **AOR of obesity (95% CI)** | ***P* value** | **AOR of obesity (95% CI)** | ***P* value** | **AOR of obesity (95% CI)** | ***P* value** |
| FF visits/time | 1.16 (1.12, 1.21) | <.001 | 1.17 (1.12, 1.22) | <.001 | 1.17 (1.12, 1.22) | <.001 |
| FF visits/food | 1.13 (1.10, 1.17) | <.001 | 1.14 (1.09, 1.18) | <.001 | 1.14 (1.10, 1.18) | <.001 |
| FF intake frequency | | | | | | |
| Infrequent | 1.06 (0.85, 1.33) | .621 | 1.12 (0.88, 1.43) | .361 | 1.13 (0.89, 1.44) | .317 |
| Moderate | 1.26 (1.03, 1.55) | .028 | 1.30 (1.05, 1.63) | .019 | 1.34 (1.07, 1.67) | .011 |
| Frequent | 1.63 (1.34, 1.99) | <.001 | 1.64 (1.32, 2.03) | <.001 | 1.75 (1.41, 2.17) | <.001 |

Abbreviations: AOR, adjusted odds ratio; FF, fast food.
[a] Binary logistic regression models for obesity adjusted for age group, gender, race/ethnicity, educational level, and income level. Sensitivity tests fit regression models to the analytic sample subtracting out respondents living in outlier census tracts demonstrating the largest change in demographic variables between 2011 and 2017 according to methods 1 and 2. Regression model results fit to the full analytic sample (full sample) are provided for comparison.
[b] Each model estimated fast food intake frequency using the fast food visit variable listed in this column as the primary independent variable.
[c] Outliers are identified by the Tukey method as values more than 1.5 times the interquartile range from each of the quartiles for a variable, i.e. upper outliers are values of the distribution > Q3 + 1.5*IQR and lower outliers are values < Q1 – 1.5*IQR.
[d] Outliers are identified as values of a variable above and below 2 standard deviations of the mean. For each method, we remove from the LACHS sample all respondents living in outlier census tracts. We then identified the union over outlier census tracts across the three measures, as some census tracts overlapped.

#### eTable 13. Adjusted Odds Ratios From Regression Models of Diabetes From Sensitivity Analyses to Time Change Between 2011-2017[a]

| | Full sample | | Method 1[c] | | Method 2[d] | |
|---|---|---|---|---|---|---|
| **Model[b]** | **AOR of diabetes (95% CI)** | ***P* value** | **AOR of diabetes (95% CI)** | ***P* value** | **AOR of diabetes (95% CI)** | ***P* value** |
| FF visits/time | 1.15 (1.08, 1.21) | <.001 | 1.17 (1.09, 1.24) | <.001 | 1.16 (1.09, 1.23) | <.001 |
| FF visits/food | 1.11 (1.06, 1.16) | <.001 | 1.12 (1.06, 1.17) | <.001 | 1.12 (1.06, 1.17) | <.001 |
| FF intake frequency (reference: never) | | | | | | |
| Infrequent | 1.08 (0.81, 1.44) | .594 | 1.12 (0.82, 1.51) | .483 | 1.12 (0.82, 1.52) | .469 |
| Moderate | 1.17 (0.89, 1.54) | .254 | 1.14 (0.86, 1.53) | .363 | 1.24 (0.93, 1.67) | .144 |
| Frequent | 1.39 (1.08, 1.81) | .013 | 1.38 (1.04, 1.83) | .025 | 1.51 (1.14, 2.01) | .004 |

Abbreviations: AOR, adjusted odds ratio; FF, fast food.
[a] Binary logistic regression models for diabetes adjusted for age group, gender, race/ethnicity, educational level, and income level. Sensitivity tests fit regression models to the analytic sample subtracting out respondents living in outlier census tracts demonstrating the largest change in demographic variables between 2011 and 2017 according to two methods (method 1 and method 2). Regression model results fit to the full analytic sample (full sample) are provided for comparison.
[b] Each model estimated FF intake frequency using theFF visit variable listed in this column as the primary independent variable.
[c] Outliers are identified by the Tukey method as values more than 1.5 times the interquartile range from each of the quartiles for a variable, i.e. upper outliers are values of the distribution > Q3 + 1.5*IQR and lower outliers are values < Q1 – 1.5*IQR.
[d] Outliers are identified as values of a variable above and below 2 standard deviations of the mean. For each method, we remove from the LACHS sample all respondents living in outlier census tracts. We then identified the union over outlier census tracts across the three measures, as some census tracts overlapped.

#### eTable 14. *P* Values for Differences Between Odds Ratios From Regression Models of Obesity and Diabetes From Sensitivity Analyses to Time Change Between 2011-2017[a]

| | ***P* value for difference[b]** | | | |
|---|---|---|---|---|
| | **Obesity** | | **Diabetes** | |
| **Model[c]** | **Method 1[d]** | **Method 2[e]** | **Method 1[d]** | **Method 2[e]** |
| FF visits/time | .81 | .81 | .73 | .86 |
| FF visits/food | .76 | .76 | .83 | .83 |
| FF intake frequency (reference: never) | | | | |
| Infrequent | .76 | .73 | .88 | .88 |
| Moderate | .87 | .76 | .91 | .82 |
| Frequent | .97 | .77 | .98 | .78 |

Abbreviations: FF, fast food.
[a] Binary logistic regression models for obesity and diabetes adjusted for age group, gender, race/ethnicity, educational level, and income level.
[b] *P* values are calculated using a $\chi^2$ test to test for the significance of difference between odds ratios from multinomial regression models fit to the full analytic sample and from regression models fit to the analytic sample subtracting out outlier census tracts demonstrating the largest change in demographic variables between 2011 and 2017 according to method 1 and 2.
[c] Each model estimated FF intake frequency using the FF visit variable listed in this column as the primary independent variable.
[d] Outliers are identified by the Tukey method as values more than 1.5 times the interquartile range from each of the quartiles for a variable, i.e. upper outliers are values of the distribution > Q3 + 1.5*IQR and lower outliers are values < Q1 – 1.5*IQR.
[e] Outliers are identified as values of a variable above and below 2 standard deviations of the mean. For each method, we remove from the LACHS sample all respondents living in outlier census tracts. We then identified the union over outlier census tracts across the three measures, as some census tracts overlapped.

**eFigure 1. Frequency distribution of the accuracy of all pings generated by a random sample of 2,000 (0.8%) of smartphone users in the mobility dataset**

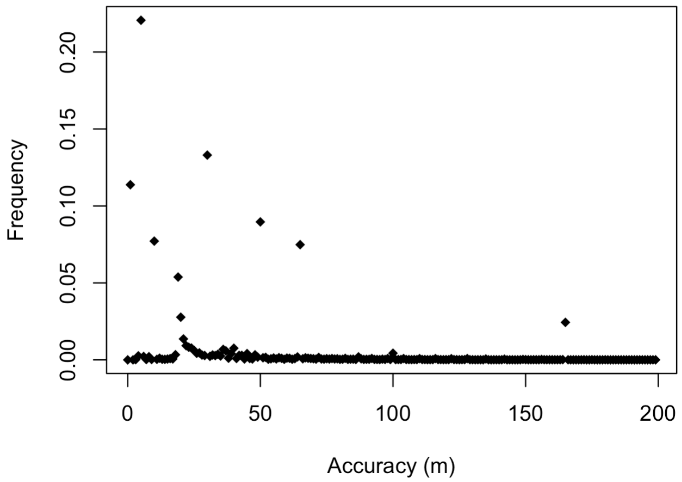


Abbreviation: meters, m.
Accuracy represents a specific level of ping GPS horizontal positioning accuracy in m. Frequency represents the share of the distribution that occurs at a specific level of accuracy. An accuracy of 0 means that the accuracy level of the ping is less than 1 m. Pings with accuracy > 200m were not included in analysis for this study.

**eFigure 2. Map of the spatial boundaries of the 272 Los Angeles County neighborhoods**

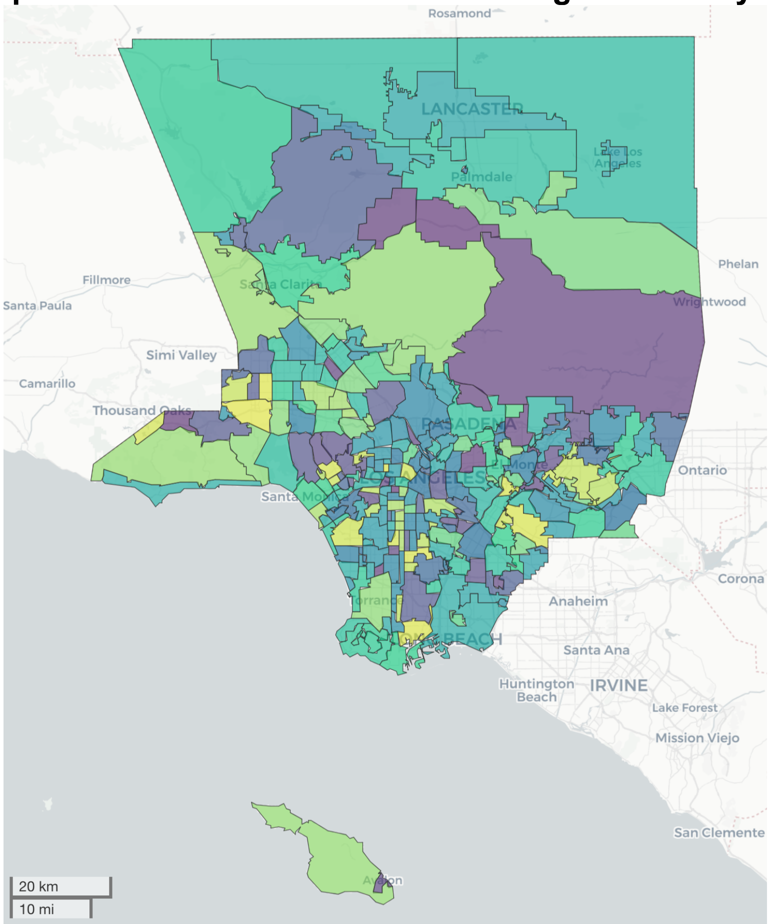

**eFigure 3. Pearson correlation between the values of each FF outlet visit variable at each value of thresholding distance $d^{max}$ between a stay and POI**

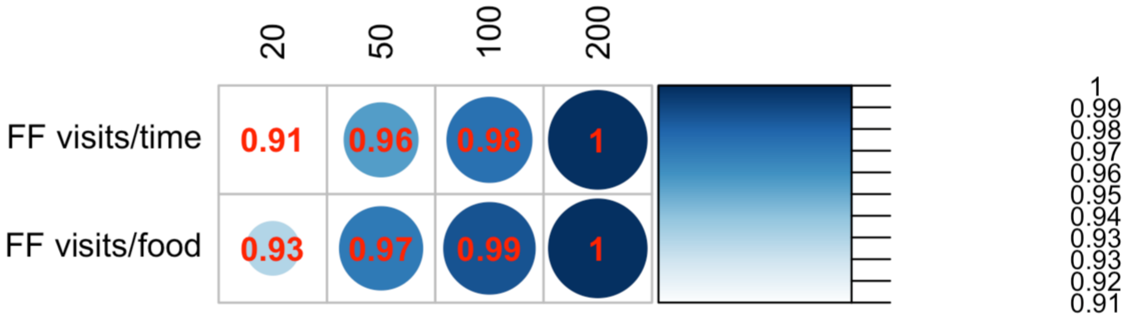


The figure presents a visual representation of the Pearson correlation coefficient between the values of the FF outlet visit variables ac LAC neighborhoods calculated at $d^{max}$=200m with that variable calculated at $d^{max}$=20, 50, and 100m. Results in the main paper are calculated at a distance of 200m.

**eFigure 4. Distribution of the Number of Smartphone Users in Analytic Sample Within Each Area for the Census Tract and neighborhood Spatial Levels**

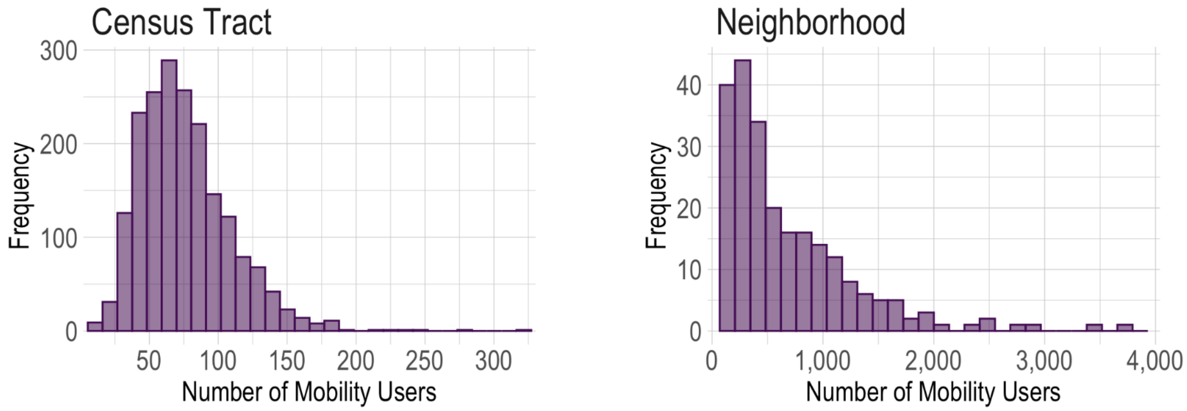

**eFigure 5: Correlation Between the Size of the Smartphone User Population Detected in Mobility Data and the Size of the Census Population at the Census Tract vs. Neighborhood Levels**

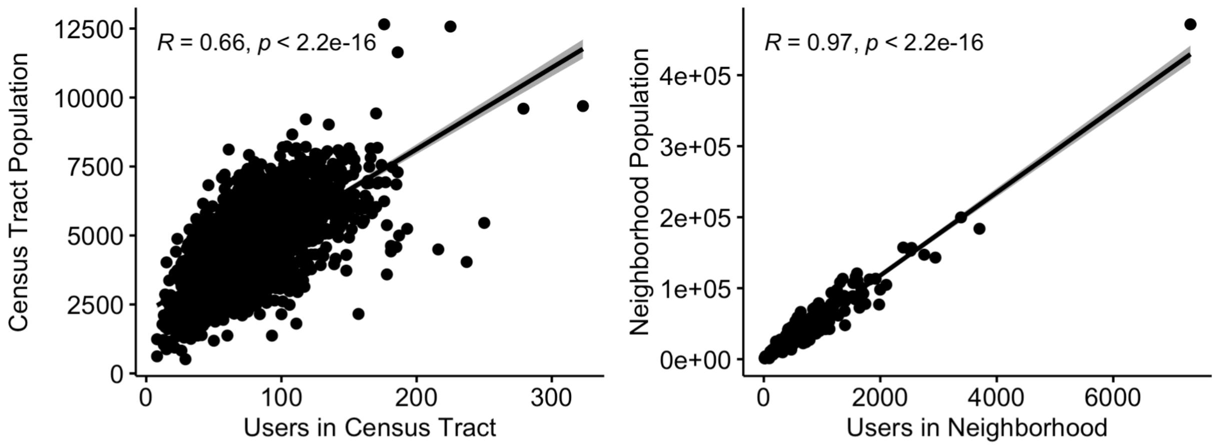


(a) Correlation of the size of the user population in each census tract detected in our smartphone data vs. size of the census population from the 2012-2016 ACS estimates (Left figure). (b) Correlation of the size of the user population in each neighborhood detected in our smartphone data vs. size of the census population from the 2012-2016 ACS estimates (Right figure).

**eFigure 6. Correlation Between Post-Stratified (Weighted) and Unweighted Fast Food Outlet Visit Variables**

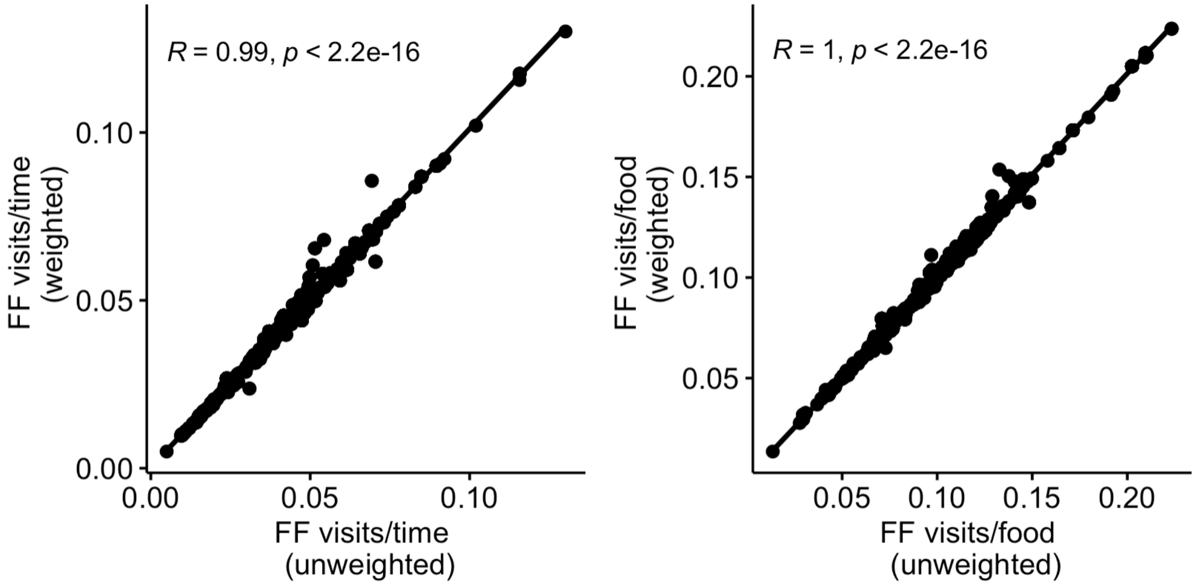


(a) Correlation of FF visits/time using the post-stratified (i.e., weighted) and unweighted values at the neighborhood level (Left figure). (b) Correlation of FF visits/food using the weighted and unweighted values at the neighborhood level (Right figure).

**eFigure 7. Histograms of the Distribution of the Unscaled Mobility Variables Linked to LACHS Respondents**

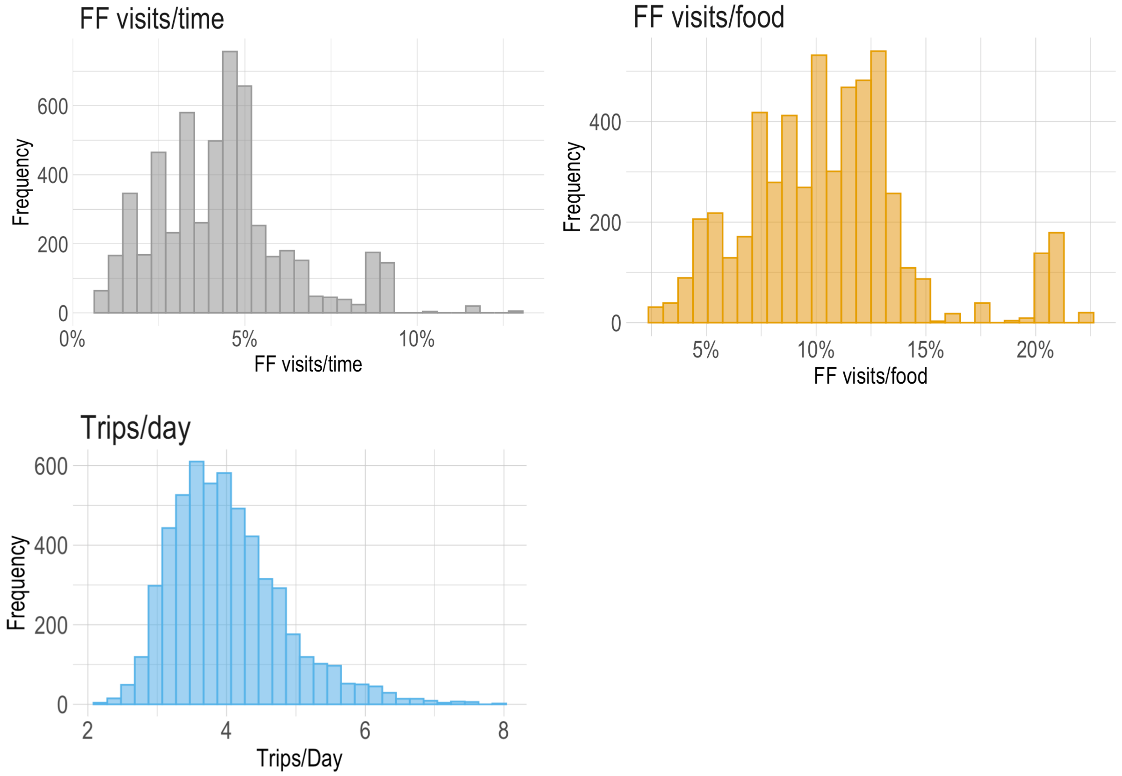


Histograms of the three mobility variables linked to LACHS respondents, before scaling from [0,10] (i.e., unscaled). The range of variable values are presented in the x-axis. Frequency counts in the y-axis represent the number of LACHS respondents with a linked mobility variable at this binned interval.

**eFigure 8. Histograms of the Differences in Three Census Tract Variables Between 2011 and 2017**

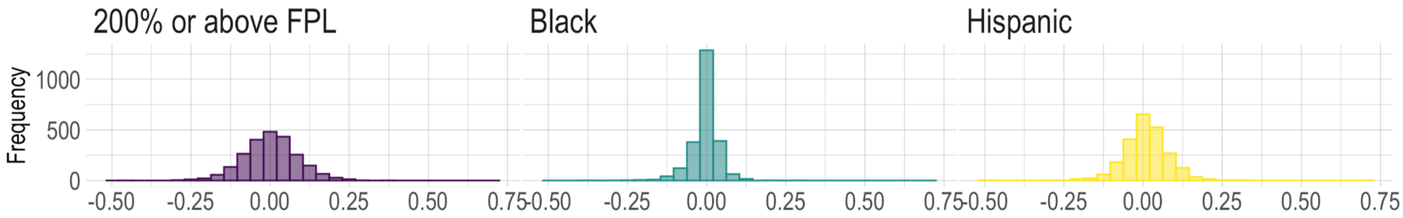


Histograms show the difference in the percent of the population in each LAC census tract above 200% of the FPL, that is Black, and that is Hispanic between 2011 and 2017.

#### eReferences


1. National Institutes of Health. The practical guide: identification, evaluation, and treatment of overweight and obesity in adults. *US Dep Heal Hum …*. Published online 2000.
2. Moro E, Calacci D, Dong X, Pentland A. Mobility patterns are associated with experienced income segregation in large US cities. *Nat Commun*. 2021;12(1). doi:10.1038/s41467-021-24899-8
3. Aleta A, Martín-Corral D, Pastore y Piontti A, et al. Modelling the impact of testing, contact tracing and household quarantine on second waves of COVID-19. *Nat Hum Behav*. 2020;4(9). doi:10.1038/s41562-020-0931-9
4. Cuebiq Data for Good Program. Accessed October 20, 2021. https://www.cuebiq.com/about/data-for-good/
5. Android Developer Reference: Location. Accessed October 20, 2021. https://developer.android.com/reference/android/location/Location
6. Apple Developer: Horizontal Accuracy. Accessed October 21, 2021. https://developer.apple.com/documentation/corelocation/cllocation/1423599-horizontalaccuracy
7. Merry K, Bettinger P. Smartphone GPS accuracy study in an urban environment. *PLoS One*. 2019;14(7). doi:10.1371/journal.pone.0219890
8. Modsching M, Kramer R, ten Hagen K. Field trial on GPS Accuracy in a medium size city: the influence of built-up. *3rd Work Positioning, Navig Commun 2006*. 2006;2006.
9. Hariharan R, Toyama K. Project Lachesis: Parsing and Modeling Location Histories. *Lect Notes Comput Sci (including Subser Lect Notes Artif Intell Lect Notes Bioinformatics)*. 2004;3234:106-124. doi:10.1007/978-3-540-30231-5_8
10. Cuttone A, Larsen JE, Lehmann S. Inferring human mobility from sparse low accuracy mobile sensing data. In: *UbiComp 2014 - Adjunct Proceedings of the 2014 ACM International Joint Conference on Pervasive and Ubiquitous Computing*. ; 2014. doi:10.1145/2638728.2641283
11. Foursquare API. https://developer.foursquare.com/
12. Choudhury SR. Foursquare pioneered the trend of “checking-in” to a place — now it sells access to its data to companies, CNBC. Published August . https://www.cnbc.com/2017/08/30/foursquare-pioneered-the-trend-of-checking-in-to-a-place--now-it-sells-your-data-to-companies.html
13. Foursquare Places. Accessed October 20, 2021. https://foursquare.com/products/places
14. Hochmair HH, Juhász L, Cvetojevic S. Data quality of points of interest in selected mapping and social media platforms. In: *Lecture Notes in Geoinformation and Cartography*. Vol 0. ; 2018. doi:10.1007/978-3-319-71470-7_15
15. Datar A, Nicosia N. Assessing social contagion in body mass index, overweight, and obesity using a natural experiment. *JAMA Pediatr*. 2018;172(3). doi:10.1001/jamapediatrics.2017.4882
16. Datar A, Mahler A, Nicosia N. Association of Exposure to Communities With High Obesity With Body Type Norms and Obesity Risk Among Teenagers. *JAMA Netw open*. 2020;3(3). doi:10.1001/jamanetworkopen.2020.0846
17. Los Angeles County Department of Public Health. County of Los Angeles Restaurant and Market Inventory. Accessed October 20, 2021. https://data.lacounty.gov/Health/COUNTY-OF-LOS-ANGELES-RESTAURANT-AND-MARKET-INVENT/jf4j-8it9
18. Fleischhacker SE, Evenson KR, Sharkey J, Pitts SBJ, Rodriguez DA. Validity of secondary retail food outlet data: A systematic review. *Am J Prev Med*. 2013;45(4). doi:10.1016/j.amepre.2013.06.009
19. The Los Angeles Times Datadesk. Mapping L.A. Neighborhoods. Accessed October 20, 2021. http://maps.latimes.com/neighborhoods/
20. United States Department of Agriculture Economic Research Service. Rural-Urban Commuting Area Codes (Updated 7/3/2019). Published 2014. Accessed October 20, 2021. https://www.ers.usda.gov/data-products/rural-urban-commuting-area-codes.aspx
21. United States Census Bureau. 2017 American Community Survey 5-Year Data. Accessed October 20, 2021. https://www.census.gov/ programs-surveys/acs
22. Salganik M. *Bit by Bit: Social Research in the Digital Age*. Princeton University Press; 2019.
23. Jiang S, Yang Y, Gupta S, Veneziano D, Athavale S, González MC. The TimeGeo modeling framework for urban motility without travel surveys. *Proc Natl Acad Sci U S A*. 2016;113(37). doi:10.1073/pnas.1524261113
24. Stern A, Narayanan A, Brown S, MacDonald G, Ford L, Ashley S. *Ethics and Empathy in Using Imputation to Disaggregate Data for Racial Equity, A Case Study Imputing Credit Bureau Data*.; 2021. https://www.urban.org/research/publication/ethics-and-empathy-using-imputation-disaggregate-data-racial-equity-case-study-imputing-credit-bureau-data/view/full_report
25. Lazer D, Hargittai E, Freelon D, et al. Meaningful measures of human society in the twenty-first century. *Nature*. 2021;595(7866). doi:10.1038/s41586-021-03660-7
